# A time-to-event heritability framework for inferring the genetic architecture of longitudinal traits

**DOI:** 10.64898/2026.02.16.26346285

**Authors:** Kodi Taraszka, Sriram Sankararaman, Alexander Gusev

## Abstract

Biobanks with longitudinal measurements have advanced our understanding of time-to-event (TTE) traits including age-of-onset and disease progression. However, limited work has characterized the heritability of TTE traits, a key parameter for comparisons of total association and predictive power. Here, we present COXMM, a Cox proportional hazard mixed model for estimating TTE heritability. Simulations show our model achieves nearly unbiased results, whereas non-TTE approaches severely underestimate TTE heritability. COXMM estimates also predict the expected accuracy of polygenic scores in survival analyses, informing study design. We analyzed a wide variety of traits and observed heritability patterns reflecting a mixture of TTE and case-control architectures with supporting GWAS and polygenic risk score analyses. Evaluating 18 trait pairs, progression to a severe condition has consistently lower heritability than all-cause incidence, suggesting that cause-specific progression may have stronger environmental influences. COXMM offers a novel framework for analyzing longitudinal disease architecture with implications for disease prediction.

## Introduction

Heritability is a fundamental parameter in genetic analyses, quantifying the fraction of trait variance that can be predicted from genetic variables as well as the expected response to selection. For decades, models for estimating the molecular heritability of common traits have primarily focused on binary and quantitative outcomes. These models rely on the same underlying phenotypic generative model (PGM)^1-4^, which assumes a linear relationship between genetic variation and the trait (for binary traits, the linear effect is on the *unobserved* liability) as well as the assumption that case-control status is invariant^2^. This PGM may be incomplete for two primary reasons. First, some individuals may be lost to follow-up prior to their disease diagnosis and therefore inaccurately classified as controls. Second, genetic variation may explain a substantial proportion of variance in the age-of-onset, which is not captured by binary phenotype models. In both cases, conventional methods would be expected to underestimate the trait heritability.

An alternative PGM treats outcomes as a time-to-event (TTE) process where genetic variation influences the instantaneous risk, or hazard, of the condition^5-7^. Under the TTE PGM and with infinite time, all individuals develop the trait; in practice, only a subset of the cases are observed due to censoring (e.g. death or loss to follow-up in the study). Classically, data of this form is analyzed with a Cox proportional hazard regression, a semi-parametric method that can model whether a genetic variant increases or decreases the relative hazard of having an event per unit of time^8^. The Cox proportional hazard model has been implemented for genome-wide association studies (GWAS), including as a mixed model, and has been shown to increase the discovery power of GWAS for TTE traits over linear models^5-7^. Although these advances are promising, the total variance in disease susceptibility attributed to all genotyped variants (i.e. SNP-heritability or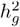) of TTE traits has remained largely unstudied. In fact, though there are numerous heritability methods for quantitative and binary phenotypes, limited work has explored the heritability of TTE traits^1-4,9,10^. This has limited the characterization of age-of-onset, disease progression, and treatment response phenotypes with previous work indicating disease progression has a distinct genetic architecture from disease susceptibility^11^. While interval phenotypes such as disease progression and treatment response are of high clinical interest, they have so far been neglected by traditional genetic studies due to a substantial loss to follow-up.

In this paper, we introduce a method for accurate heritability estimation of longitudinal/TTE traits. Our method, COXMM, is a Cox proportional hazard Mixed Model which accounts for both age-of-onset and censoring. We extensively evaluate COXMM under various PGMs, both for TTE traits and case-control traits. COXMM provided accurate estimates of heritability across TTE simulation frameworks while conventional methods applied to the case-control status and (case-only) age-of-onset provided greatly deflated estimates, indicating that the right PGM was vital for accurate heritability estimation. Using longitudinal data from the UK Biobank, we applied COXMM to seven cardiovascular traits, 18 disease progression phenotypes between three cardiometabolic risk traits and other cardiovascular events, and nine additional traits across a variety of disease categories with age-of-onset and diagnosis status readily available. Overall, our work shows COXMM to be an effective method for estimating the heritability of TTE traits and demonstrates that the genetic architecture of many traits may be a composite of the TTE PGM and LTM PGM.

## Results

### COXMM provides empirically unbiased heritability estimates for time-to-event traits

We propose COXMM, a method for estimating molecular heritability under a time-to-event (TTE) phenotypic generative model (PGM). When defining a PGM for any trait, an underlying assumption is that trait variability can be separated into two additive variance components: a genetic component and all other effects, which we broadly refer to as the “environment” (**Fig. 1A**). Under the classic liability threshold model (LTM), the genetic effect (in conjunction with the environment) invariantly determines if the trait ever develops by influencing the *unobserved* liability: individuals above the liability threshold are effectively lifetime cases at birth and individuals below the threshold are effectively lifetime controls. The LTM PGM has previously been linked to the age-of-onset for cases by imposing earlier onset on individuals with higher liability^12^. In contrast, the TTE PGM assumes that over an infinite timespan everyone develops the trait as a function of the baseline hazard (indirectly capturing environmental effects) and the genetic effects on the hazard (or hazard ratios). Under the TTE generative model, the LTM “controls” are simply unobserved cases due to censoring (**Fig. 1B**).

**Fig 1.**
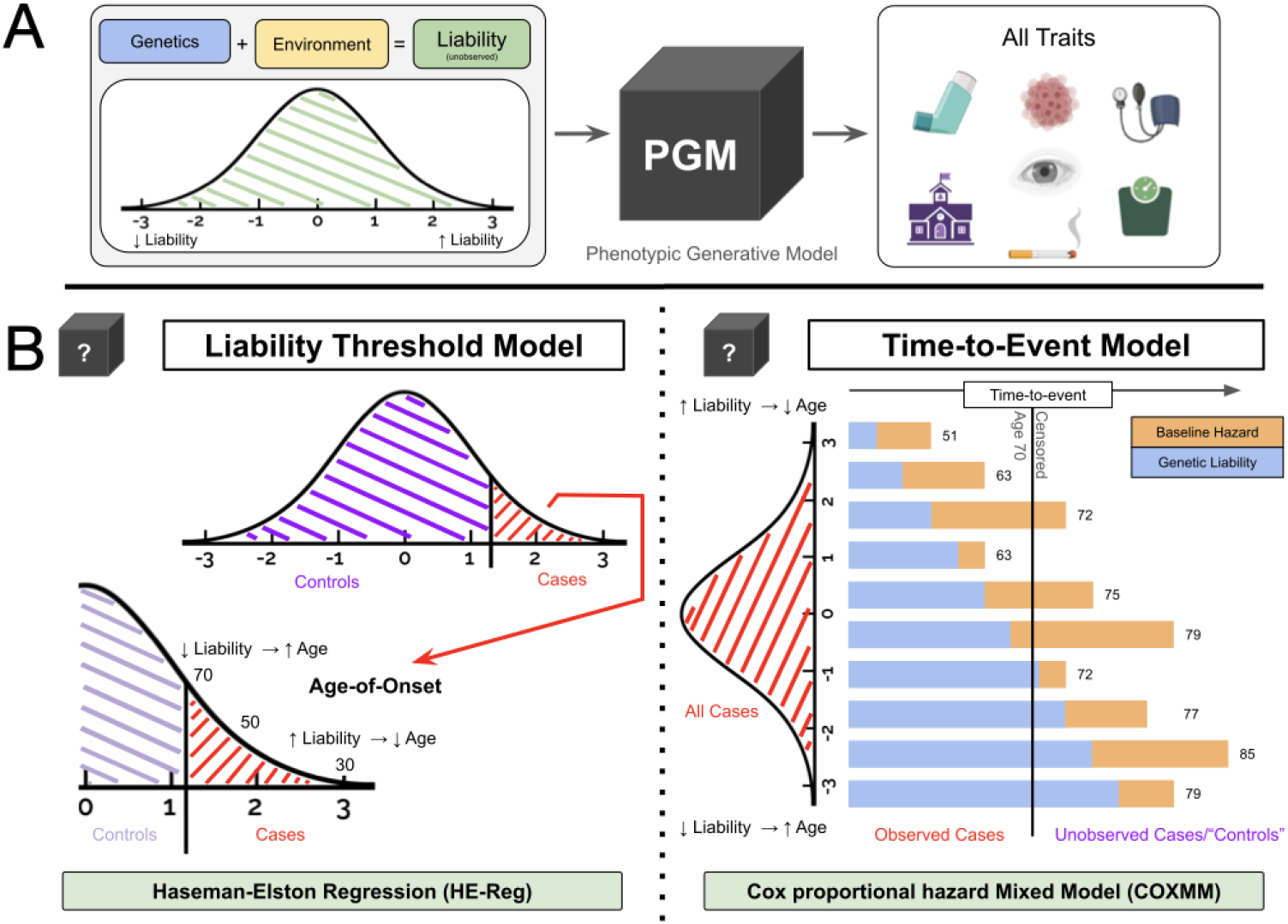
COXMM Overview. **A**. Genetics and environment (i.e. all non-genetic effects) contribute to the phenotypic generative model (PGM) for all traits. **B**. Traits which are truly case-control are generated under the Liability Threshold Model (LTM; left) while time-to-event (TTE) traits are generated by the TTE model (right).

To explicitly define the TTE PGM, let *X*_*N* × *M*_ be the standardized genotype matrix measured for *N* individuals across *M* SNPs. Further, let *λ* (*t*|*X*) be a *N* × 1 vector of their hazard at time *t* such that:

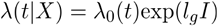

where *λ* _0_(*t*) is the unspecified baseline hazard,*l*_*g*_ is the genetic liability (random effect), and *I* is the identity matrix. We assume the genetic liability, 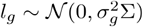, follows the Gaussian distribution centered at zero with the variance determined by the the genetic variance component 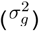 and genetic relatedness between individuals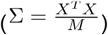. COXMM estimates 2 the 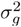 parameter by iteratively solving a penalized partial likelihood and the integrated marginal likelihood defined by a Laplace approximation until convergence^13,14^. We note that there is no explicit modeling of an environment term which is assumed to be captured within the unspecified baseline hazard. Any ties in the TTE are addressed using Breslow’s approximation, and the standard errors are estimated using a weighted block jackknife^15-17^. Finally, we transform the 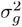 estimate into a heritability estimate defined on the log frailty scale. This transformation is based on previous work showing that the log of the conditional integrated hazard function (which measures the accumulated risk of an event) follows the extreme value distribution and is equivalent to a linear model on the log cumulative baseline hazard scale (log(Λ_0_(*T*_*i*_))^18^The unconditional variance of log(Λ_0_(*T*_*i*_) is Var(*l*_*gi*_) + π^2^/6 from which we can then define the proportion of accumulated hazard attributed to genetics or heritability to be 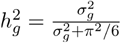 (see **Methods**)^18^.

We assessed the accuracy of COXMM estimates under this TTE PGM by simulating data under a Weibull distribution with a shape parameter of 1 and the scale parameter set by *l*_*g*_ which is generated using simulated common SNPs whose effect sizes follow the infinitesimal model (See Methods). The censoring event was assumed to be a cohort censoring where the individuals with an onset beyond a threshold were unobserved/controls. COXMM provided an empirically 2 unbiased estimate of 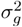 across the evaluated parameter range (**Fig 2A**; **Supplementary Table 1**). COXMM was empirically unbiased when *K* ∈ {0.2, 0.4}for all values of 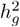 though COXMM became slightly deflated at higher values 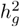 of when K was very low (*K* =0.05; **Fig 2A-B**; **Supplementary Table 1**). For the same simulations, we compared COXMM 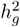 estimates to two conventional frameworks: (i) treating individuals that experienced an event prior to censoring as cases and the rest as controls, followed by a conversion to the liability-scale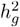; or (ii) restricting to cases and treating their age-of-onset as a continuous phenotype to which we applied a rank-inverse normal transformation. Haseman-Elston regression (HE-Reg), a classic heritability estimator that is robust to case-control ascertainment, was applied for both trait definitions. Both age-of-onset HE-Reg and case-control HE-Reg were severely downward biased across all configuration of 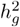 and when *K* ∈ {0.05,0.2}. For example, when the true 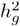 was 0.33, the case-control HE-Reg estimate (averaged over 50 simulations) was 0.17, nearly half the true parameter value; age-of-onset estimate was even further deflated with an averaged estimate of 0.007 (**Fig 2B**; **Supplementary Table 1**). Interestly, when *K =* 0.4 (i.e the proportion of cases is high), age-of-onset HE-Reg remained downward biased but case-control HE-Reg was nearly unbiased and was even slightly inflated for higher values of 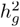 (**Fig 2B**; **Supplementary Table 1**). In sum, our simulations showed that under a TTE PGM, COXMM was nearly unbiased and consistently produced a more accurate heritability estimate than case-control HE-Reg and age-of-onset HE-Reg, particularly when the overall (observed) trait prevalence is low.

**Fig 2.**
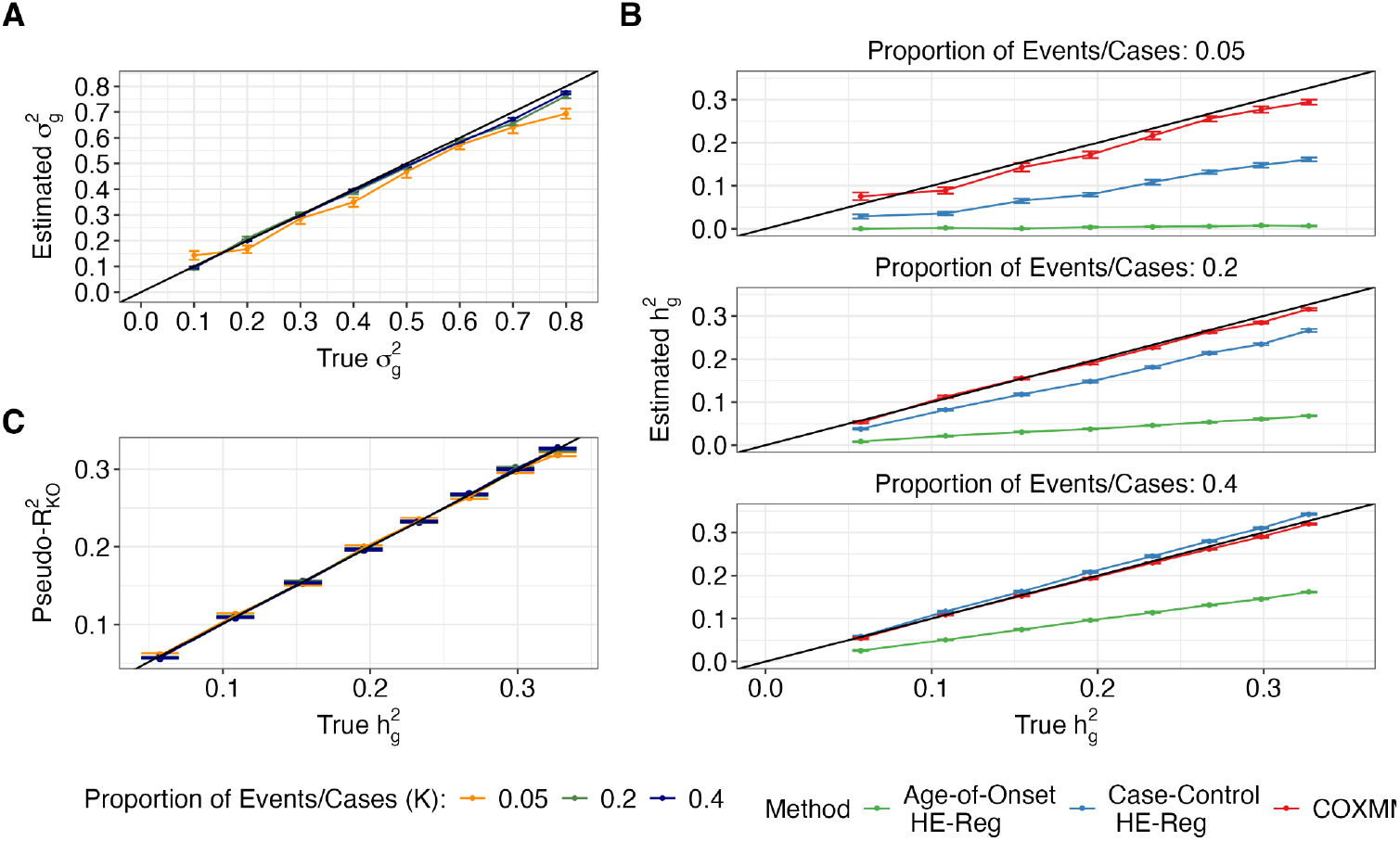
COXMM Simulations. We simulated TTE traits using a Weibull distribution with the shape parameter set to 1 and the scale parameter set using genetic variance component values 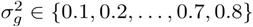 and aggregate the estimates for each proportion of observed events/cases (*K* ∈ {0.05,0.2,0.4}). We reported the mean 2 parameter estimate and its standard error over 50 simulations. **A**. The x-axis corresponded to the true 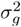 parameter and the y-axis reflected the estimated 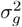 for COXMM with a black line indicating the ground truth with the colors corresponding to different proportions of observed events/cases. **B**. For the same simulations, we reported the heritability 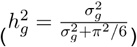. The x-axis corresponded to the true heritability and the y-axis reflected the estimated heritability. We split the results into sub-panels based on proportion of observed events/cases (*K* ∈ {0.05,0.2,0.4}). The results for COXMM were shown in red, age-of-onset HE-Reg in green, and case-control HE-Reg in blue. **C**. The x-axis is the true heritability 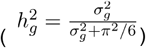. The y-axis reflects the average model fit metric based on the pseudo-R^2^ defined by Kent and O’Quigley with a black line reflecting the linear model fit between 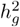 and pseudo-R^2^.

As SNP-heritability estimates are commonly used to forecast the expected upper bound on predictive accuracy (R^2^) of the Best Linear Unbiased Predictor (BLUP) or optimal linear polygenic score, we sought to investigate how our estimated parameters 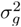 and 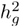tracked with predictive accuracy for a TTE model. Notably, a variety of metrics exist for quantifying the predictive accuracy of a TTE model, including various flavors of concordance, d-statistics, and pseudo-R^2 19-22^. In simulated data with an optimal/noise-free genetic predictor, we found a linear relationship between COXMM 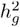 and four metrics of TTE fit: concordance, Royston and Sauerbrei D-statistic, Kent and O’Quigley’s pseudo-R^2^, and Nagelkerke’s pseudo-R^2^ (**Fig 2C**; **Supplementary Fig 1**; **Supplementary Table 2**)^19-22^. In particular, the COXMM 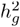 can be interpreted as the approximate upper bound on Kent and O’Quigley’s pseudo-R^2^ from a Cox regression with an optimal genetic predictor (**Fig 2C**). As a result, we report COXMM 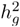 parameters in all subsequent analyses unless otherwise noted.

### COXMM is robust to modifications to the TTE PGM

We next simulated traits under a number of alternative TTE frameworks to assess the robustness of COXMM to model misspecification. First, we evaluated the impact of covariates such as sex and age as well as different censoring events, following previous work^5-7^. COXMM was robust to the inclusion of fixed effects (**Fig. 3A**; **Supplementary Table 1**) and the censoring process (**Supplementary Table 1**). Second, we simulated TTE data under other plausible Weibull distributions. COXMM was robust to the Weibull shape parameter as well (**Fig. 3B**; **Supplementary Table 1**). Overall, COXMM was stable across TTE PGMs except when *K* was very low (*K* = 0.01; **Supplementary Fig 2**; **Supplementary Table 1**).

**Fig 3.**
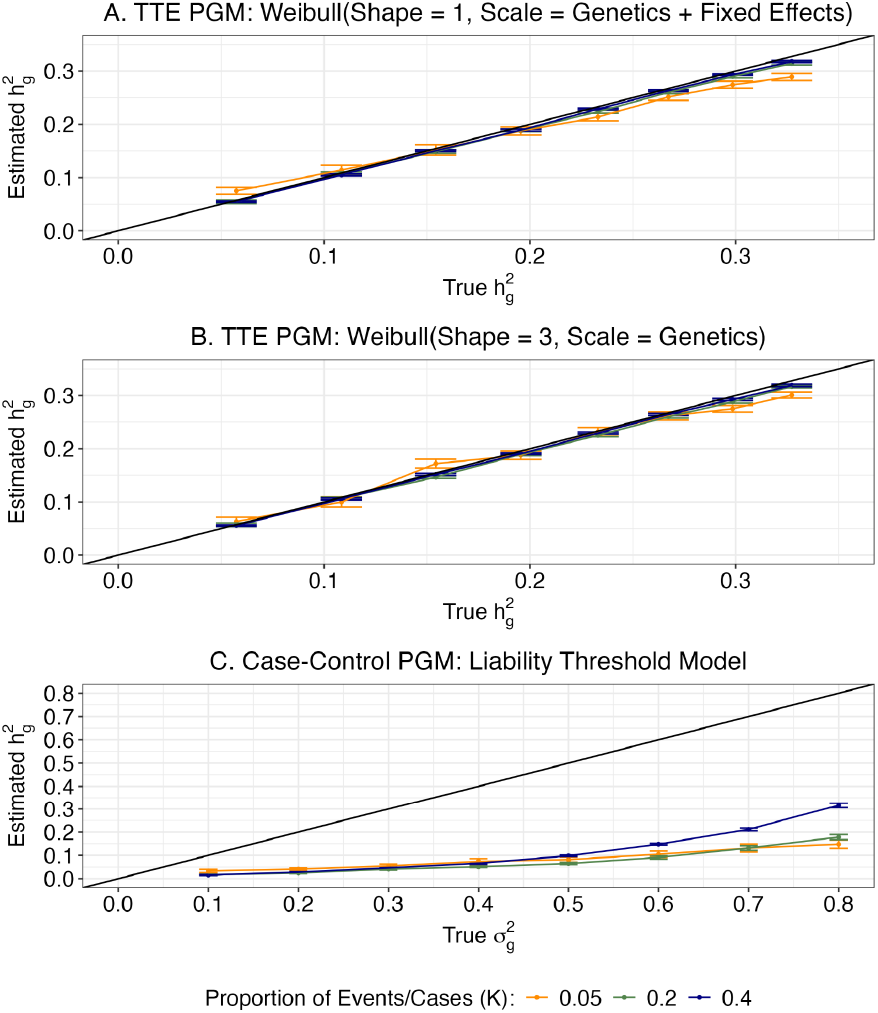
COXMM performance under various phenotypic generative models (PGMs) In each panel, we simulated a different phenotypic generative model (PGM) across genetic variance component values: 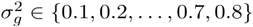 and proportion of observed events/cases (*K* ∈ {0.05, 0.2,0.4}). Within panels A and B, the x-axis corresponded to the true heritability 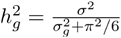 while in panel C the x-axis corresponds to 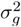. In all panels, the y-axis reflected the estimated heritability with a black line indicating the ground truth. **A**. We simulated 50 traits under the time-to-event (TTE) PGM using a Weibull distribution with the shape parameter set to 1 and the scale parameter accounting for the genetic liability and two fixed effect variables with the total fixed effect variance set to 0.5. **B**. We simulated 50 traits under the TTE PGM using a Weibull distribution with a shape parameter of 3 and the scale parameter only based on the genetic liability. **C**. We simulated 50 traits under the case-control PGM using the classic LTM and note the x-axis is in terms of 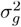 (which is equivalent to 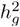 under the LTM).

We additionally investigated how COXMM performs when the generative model was grossly misspecified by simulating data under the case-control PGM (i.e LTM; **Fig 3C**). As expected, case-control HE-Reg produced unbiased heritability estimates across all simulations (**Supplementary Table 1**)^23^. In order to generate a TTE trait, we used a liability induced age-of-onset model and set the age-at-censoring in a similar fashion (see Methods). COXMM consistently reported downward biased heritability estimates under this PGM due to the mismatch in generative models (**Fig. 3C**; **Supplementary Table 1**). For example, when the true ^*h*^*-g* was 0.3 under the LTM, the COXMM estimate (averaged over 50 simulations) was 0.12 which is less than half of the ground truth value. Finally, we evaluated case-only age-of-onset HE-Reg across the various models as age-of-onset heritability measurements have recently been reported^23-25^. Age-of-onset HE-Reg consistently reported deflated heritability estimates regardless of the PGM, LTM and TTE (**Supplementary Table 1**).

Overall these simulations demonstrate that heritability is consistently estimated more accurately when the inferential method matches the generative PGM and that deflated estimates are reported when the PGM is misspecified. Comparisons of TTE and LTM heritability estimates for the same trait can thus provide evidence in support of one disease model over the other, particularly when the trait prevalence is low and the model estimates are expected to diverge substantially.

### Genetics influence both the age-of-onset and case-control status of cardiovascular traits

We applied COXMM to real data using seven well-measured cardiovascular traits in the UK Biobank generated from the ukpheno R package (**Supplementary Table 3**)^26^. Our selected traits were a composite of self-reported data, ICD codes, medications, hospital records, and they contained both the diagnosis status and age-at-diagnosis (or censoring). All traits had an observed prevalence of at least 4% (**Supplementary Table 3**). Surprisingly, the mean heritability estimates were statistically indistinguishable between the three methods: COXMM, case-control HE-Reg, and case-only age-of-onset HE-Reg (mean estimate across traits: COXMM = 0.20, case-control HE-Reg = 0.23, and age-of-onset HE-Reg = 0.15; COXMM and case-control HE-Reg Welch’s t-test p = 0.48; COXMM and age-of-onset Welch’s t-test p = 0.16; **Fig. 4A**; **Supplementary Table 3**). These findings do not align with our simulations under the canonical PGMs and suggest a more complex genetic architecture (**Fig 2**; **Fig 3**; **Supplementary Table 1**; **Supplementary Table 3**).

**Fig 4.**
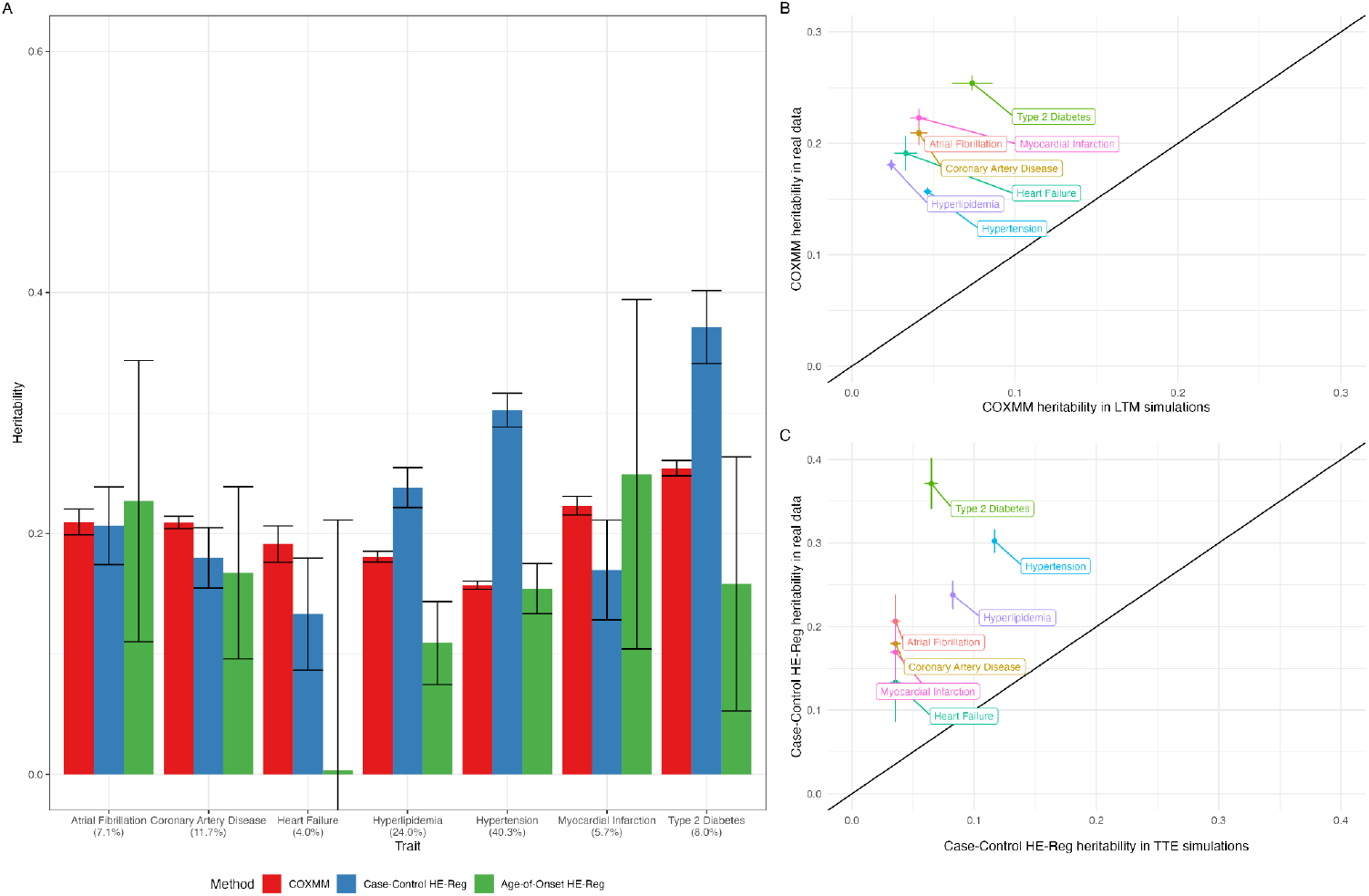
Comparison of heritability estimators using seven cardiovascular traits. **A**. We estimated the heritability of seven age-at-diagnosis traits measured in the UK Biobank using three approaches. The traits are separately reported along the x-axis with the case prevalence reported under each trait. A bar graph depicts the estimated heritability and standard error for each method along the y-axis: case-control HE-Reg in blue, case-only age-of-onset HE-Reg in green, and COXMM in red. **B**. We compared the COXMM heritability estimates for the cardiovascular traits (y-axis) to the COXMM heritability estimate under the LTM PGM simulations. Each simulation’s heritability estimate for COXMM was chosen based on the ground true heritability parameter that was closest in absolute value to the Case-Control HE-Reg real data estimate and the proportion of cases/K closest in absolute value to the disease prevalence in the UK Biobank. **C**. We compared the Case-Control HE-Reg heritability estimates for the cardiovascular traits (y-axis) to the Case-Control HE-Reg heritability estimate under the TTE PGM simulations. Each simulation’s heritability estimate for Case-Control HE-Reg was chosen based on the ground true heritability parameter that was closest in absolute value to the COXMM real data estimate and the proportion of cases/K closest in absolute value to the disease prevalence in the UK Biobank.

To further investigate the underlying PGM, we compared the COXMM and case-control HE-Reg estimates for each trait as these produced respectively unbiased heritability estimates in our simulations (**Fig 4A**; **Supplementary Table 3**). For 3/7 traits (hypertension, hyperlipidemia, and type 2 diabetes), the case-control HE-Reg estimate was significantly higher than COXMM (Wald’s test p < 7.14 x 10^−3^; **Supplementary Table 3**). However, the COXMM estimate for all three traits was also significantly higher than would be expected under a pure LTM model (**Fig 4B**; **Supplementary Fig 2**). For the remaining four traits, the COXMM heritability point estimate was higher than case-control HE-Reg though none of the differences were nominally significant. However, the LTM estimate for all four traits was significantly higher than would be expected under a pure TTE model (**Fig 4C**). We additionally tested the influence of age-at-enrollment as a covariate and did not observe a systematic difference for COXMM and, therefore, did not include the covariate throughout. (**Supplementary Fig 7**; **Supplementary Table 3**). In sum, the real data results for these cardiovascular traits did not reflect either a pure TTE PGM or a pure LTM PGM, suggesting that the genetic architecture is instead a complex combination of the two PGMs.

We next tested the robustness of our heritability estimates through a series of sensitivity analyses. We focused on hypertension as the test trait due to its high prevalence (*K* = 0.4; **Supplementary Table 3**). First, we right censored hypertension such that cohort censoring events occurred at a specified age for seven thresholds: 45, 50, 55, 60, 65, 70, and 75 (**Supplementary Fig 5**; **Supplementary Table 3**). For case-control HE-Reg, all events under the specified age were set to cases and all other individuals were treated as controls reflecting the common limitation of loss to follow-up for the LTM framework. COXMM estimates were robust across censoring ages while case-control HE-Reg produced an upward biased estimate of the lifecourse heritability when using the population prevalence of events when transforming to the liability-scale heritability; *K* = 0.4; **Supplementary Fig 4; Supplementary Table 3**). We note that this bias may be mitigated by using the in-sample prevalence as the true population prevalence but such an assumption does not follow standard conventions and is unlikely to be effective for all traits (**Supplementary Fig 5; Supplementary Table 3**).

### Cardiovascular disease progression phenotypes are heritable

We next considered a set of 18 interval/progression phenotypes, defined as the time between the respective age-at-diagnosis for three cardiometabolic risk index traits (type 2 diabetes, hypertension, and hyperlipidemia) and the remaining traits (**Fig 5A)**. For each focal trait, we restricted to individuals who developed the indexing event and then either modeled the time until the second event/loss-to-follow-up (COXMM) or the presence of the second event (case-control HE-Reg); for case-control HE-Reg, we used the observed prevalence for adjusting to the liability scale heritability. We did not evaluate age-of-onset HE-Reg due to the further reduced sample sizes for the case-only analyses (**Fig. 5**; **Supplementary Table 3**). Surprisingly, the average case-control HE-Reg heritability estimate was significantly higher than the average COXMM heritability estimate across the progression phenotypes (mean estimate across traits: COXMM = 0.12, case-control HE-Reg = 0.19; Welch’s t-test p = 0.02), and the heritability estimates from the two models were highly correlated (*ρ* = 0.9, p = 5.3×10^−7^). 2/18 traits had a significantly higher case-control HE-Reg estimate individually (Wald’s test p < 2.8×10^−3^; hypertension->hyperlipidemia and hypertension->type 2 diabetes; **Fig 5B**; **Supplementary Table 3**). COXMM identified two uniquely significant heritability estimates: the progression from type 2 diabetes to myocardial infarction and the progression from hypertension to heart failure (**Supplementary Table 3**). Notably, for the interval between hypertension and heart failure the COXMM heritability was only negligibly higher than case-control HE-Reg (a difference of 0.002) and for the interval between type 2 diabetes and myocardial infarction the COXMM estimate was actually lower than the case-control HE-Reg; however case-control HE-Reg had large standard deviations for both traits resulting in non-significant heritability estimates. Lastly, the progression from hyperlipidemia to atrial fibrillation was only associated with case-control HE-Reg (**Supplementary Table 3**). Overall, the low and largely non-significant individual estimates for COXMM may indicate truly little genetic effect on the timing of disease progression, or limited statistical power in this cohort. For those traits that did exhibit significant heritability, the indistinguishability between the two PGMs remains and again points to a complex genetic architecture.

**Fig 5.**
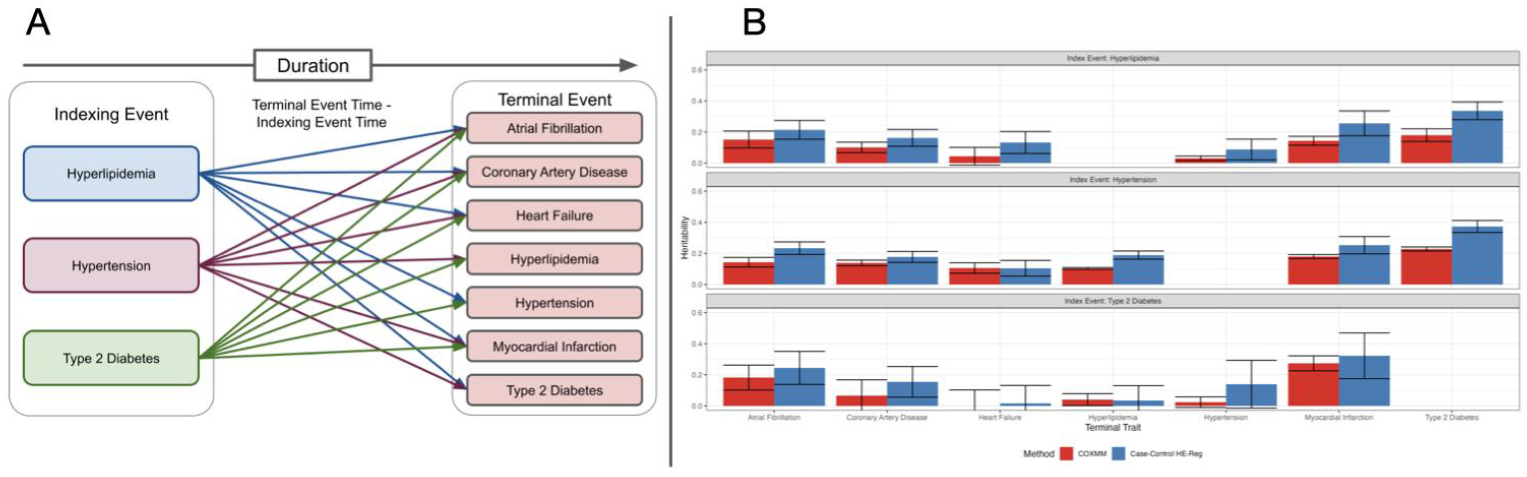
Comparison of heritability estimators using 18 disease progression traits. **A**. A schematic of the 18 disease progression traits from UK Biobank defined by the duration of time between the age-at-diagnosis for three indexing traits: hyperlipidemia, hypertension, and type 2 diabetes and the terminal traits: atrial fibrillation, coronary artery disease, heart failure, hyperlipidemia, hypertension, myocardial infarction, and type 2 diabetes. **B**. The terminal traits are separately reported along the x-axis with a bar graph depicting the estimated heritability and standard error for each method along the y-axis: case-control HE-Reg in blue and COXMM in red separated into rows based on the indexing event.

We next explored biological factors that may be driving the disease progression heritability. First, we evaluated whether the disease progression phenotype was merely an indirect measure of either the indexing or the terminal event. The heritability of disease progression was not significantly correlated with the heritability of the indexing phenotype for either method (COXMM: p = 0.32; case-control HE-Reg: p = 0.42). However, the heritability of disease progression was significantly correlated with the heritability of the terminal trait for COXMM (*ρ* = 0.78, p = 1.2×10^−4^) but, interestingly, not for case-control HE-Reg (*ρ* = 0.40, p = 0.10). While highly correlated, the heritability of the disease progression phenotype was consistently lower than the heritability of the terminal phenotype for COXMM (Welch’s t-test p = 1.6×10^−4^) while the uncorrelated case-control HE-Reg estimates were indistinguishable (Welch’s t-test p = 0.39). The significantly correlated though distinct heritability estimates for COXMM may indicate that the disease progression phenotypes identified uniquely heritable pathways for developing the terminal trait. Finally, we restricted to the 8/18 significantly non-zero case-control HE-Reg heritability estimates and observed a significant correlation between the disease progression and terminal phenotype heritabilities (*ρ* = 0.81, p = 0.01; Welch’s t-test p = 0.7), suggesting that COXMM had higher precision to identify this result across all traits. Overall, these findings indicate a strong relationship between the interval phenotype and terminal trait heritability measurements for both methods though for COXMM the significantly decreased heritability for disease progression may indicate a larger environmental influence.

We next explored whether distinct heritable pathways to the terminal trait could be explicitly identified through different index traits, focusing on CAD as the focal trait. We propose three possible models of disease progression to CAD: (1) progression to CAD through an index event is simply a random subset of all-cause progression to CAD and has comparable COXMM heritability to all-cause CAD; (2) progression to CAD through an index event is more environmentally mediated (e.g. because of therapeutic interventions on the index event) and has lower COXMM heritability than all-cause CAD, while developing CAD *without* an index event has higher COXMM heritability than all-cause CAD; (3) progression to CAD through an index event identifies a distinct disease subtype and has higher COXMM heritability than all-cause CAD. Overall, our findings were consistent with the second hypothesis that progression to CAD via an index event was more environmentally mediated than direct progression to CAD. First, restricting to individuals that developed CAD without *any* of the three indexing events produced significantly higher COXMM heritability than all-cause CAD (No Indexing Event ->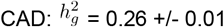; age-of-onset 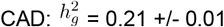; Wald’s test p = 8.7×10^−7^). Second, restricting to individuals who developed CAD without *one* of the three indexing events also produced higher COXMM heritability than all-cause CAD, though only 2/3 differences were significant (CAD without hyperlipidemia versus age-of-onset CAD, Wald’s test p = 1.4×10^−4^; CAD without hypertension versus age-of-onset CAD, Wald’s test p = 4.0×10^−3^; CAD without type 2 diabetes versus age-of-onset CAD, Wald’s test p = 0.7; **Supplementary Table 3**). In contrast, for case-control HE-Reg, heritability estimates of developing CAD without a single indexing event or without any indexing events was indistinguishable from all other estimates (**Supplementary Fig 5**; **Supplementary Table 3**). Lastly, all-cause CAD heritability was generally higher than the heritability for progression from an index event to CAD. For hypertension and CAD, the COXMM heritability for all-cause CAD was significantly higher than the COXMM heritability for progression from hypertension to CAD (Wald’s test p = 8.2×10^−5^) as well as significantly higher than the COXMM heritability estimate for the subset of CAD with hypertension (Wald’s test p = 5.9×10^−3^). For hyperlipidemia:the COXMM heritability for all-cause CAD was significantly higher than the COXMM heritability for progression from hyperlipidemia to CAD (Wald’s test p = 7.4×10^−4^), as well as significantly higher than the COXMM heritability for the subset of CAD with hyperlipidemia (Wald’s test p = 3.7×10^−4^). Lastly, the COXMM heritability for disease progression from type 2 diabetes to CAD was not significantly different from zero (COXMM: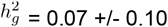) and was also indistinguishable from all-cause age-of-onset CAD (Wald’s test p = 0.08); again, the case-control HE-Reg heritability estimates were comparable to each other (one sided Wald’s test p = 0.40; **Supplementary Table 3**). Interestingly, case-control HE-Reg heritability estimates generally did not differ between all-cause CAD and index-based subsets of CAD, highlighting the unique perspective that COXMM can provide on disease progression. Overall, the higher COXMM heritability estimates for those who developed CAD without indexing event(s) and lower COXMM heritability estimates for progression from an index event to CAD supports the hypothesis that disease progression through intermediate states exhibits increased environmental variance relative to disease progression directly to the terminal phenotype (even after accounting for age-of-onset via COXMM).

We performed a number of sensitivity analyses on the disease progression heritability estimates. We tested the effect of a decreased sample size in hypertension and observed negligible changes after randomly subsampling to 50%, 25% and 10% of the cohort (**Supplementary Table 3**). We considered the effect of left truncation of the terminal trait, as the requirement of an indexing event would naturally induce a shift towards a later age-of-onset in the terminal trait. We removed all individuals whose hypertension onset (or censoring event) happened before the age of 60 and separately before the age of 70 and observed a decrease in the heritability estimate for both COXMM and case-control HE-Reg (COXMM: all samples hypertension 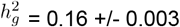; hypertension onset over 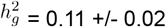; hypertension onset over 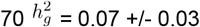; COXMM: all samples hypertension 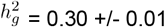; hypertension onset over 60 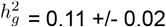; hypertension onset over 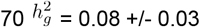; **Supplementary Table 3**). This trend is consistent with a larger genetic effect on those with a younger age-of-onset in the terminal trait. Interestingly, while left truncation produced significantly lower case-control HE-reg heritability, restricting on an index event (above) did not produce significantly lower case-control HE-reg heritability, suggesting that the consistently lower COXMM disease progression heritability cannot be explained by older age of onset / left truncation alone.

### Time-to-event and case-control polygenic risk scores were jointly predictive of cardiovascular traits

We validated the heritability estimates for the seven age-related cardiovascular traits by comparing the polygenic risk scores (PRSs) generated under the case-control PGM to those generated under the TTE PGM. We hypothesized that if the true disease architecture is a mixture of TTE and LTM, then polygenic scores learned from both models would be jointly predictive of the outcome. We separately ran the TTE GWASs and the case-control GWASs using 90% of the cohort and used the resulting association statistics to construct two respective PRSs in the held-out individuals (see Methods). For each trait, we standardized the PRSs and fit four Cox proportional hazard regression models and four logistic regression models; the models were as such: (1) a joint model with both the case-control PRS and TTE PRS (2-3) each PRS alone, and (4) a null model with only the covariates (see Methods). We then compared the AIC between the models to better understand how informative the PRSs were for the traits. When testing the PRS in a Cox proportional hazard regression, jointly modeling both the case-control PRS and TTE PRS achieved the best AIC for 3/7 traits while the TTE PRS alone had the lowest AIC for the remaining four traits (**Supplementary Table 4**). Strikingly, a similar relationship was observed when testing the PRS in a logistic regression, where jointly modeling both the case-control PRS and TTE PRS was the best model for 2/7 traits and the TTE PRS alone had the lowest AIC for the remaining five traits (**Supplementary Table 4**). Consistent with genetics influencing both TTE and case-control status, 5/14 regressions were best fit using both PRSs with the remaining 9/14 were best fit using just the TTE PRS.

We next conducted the same PRS validation analysis for the 18 disease progression traits. In the Cox proportional hazard regression, jointly modeling both the case-control PRS and TTE PRS produced the lowest AIC for 1/18 traits while the TTE PRS alone was the best model for 8/18 traits and the case-control PRS alone had the lowest AIC for 4/18 traits (**Supplementary Table 4**). The best model according to the AIC for the remaining five traits was the null model (i.e. exclude the PRSs altogether). As the AIC penalizes the inclusion of additional features and many of the heritability estimates were quite low, this may not be unexpected. When we compared the logistic regression AIC for these 18 traits, jointly modeling both the case-control PRS and TTE PRS was the best model for 3/18 traits (**Supplementary Table 4**). Modeling the TTE PRS alone was best for 2/18 traits while the case-control PRS alone produced the optimal AIC for 7/18 traits. Finally, for the remaining 6/18 traits, the null model was determined to be the best fit in the logistic regression. We note that we did not include the age-of-onset PRS in these analyses due to the limited sample size which also hindered GWAS discovery power (see below). These analyses further underscore that disease progression traits are genetically distinct from the age-at-diagnosis traits: the environmental effect was most prominent in 11/36 (null model as the best fit model: 5/18 Cox proportional hazard regressions and 6/18 logistic regressions), 14/36 models were improved by the inclusion of the TTE PRS, and the remaining eleven models reported only including the case-control PRS was the best fit model.

### Novel genetic associations are identified in disease progression traits

We next tested the validity of the heritability estimates by comparing the discovery power of three GWAS methods: TTE GWAS (SPACox), case-control GWAS (logistic regression) and a case-only age-of-onset GWAS (linear regression)^5^. For 6/7 cardiovascular traits, the TTE GWAS identified more independent loci than the other two methods indicating the TTE GWAS had more discovery power for these age-related traits (368 loci for TTE GWAS, 301 for case-control GWAS, and 31 for age-of-onset GWAS respectively; **Supplementary Table 5**). We note that the other trait, heart failure, only had one unique locus for both the TTE GWAS and case-control GWAS. In total, 76/368 loci were uniquely identified by the TTE GWAS while the case-control GWAS had 10 unique loci and the age-of-onset GWAS identified zero (**Supplementary Table 5**). We additionally conducted GWAS analyses on the 18 interval phenotypes and observed a similar phenomenon with TTE GWAS being comparable to the case-control GWAS (TTE GWAS: 65 loci with six unique to TTE; case-control GWAS: 64 loci with five unique to case-control GWAS, and age-of-onset GWAS: two loci with both being unique to age-of-onset GWAS; **Supplementary Table 5**). Overall, our findings indicated that TTE GWAS had substantially more discovery power than logistic or linear regression in the seven age-related traits and a similar discovery power in the 18 interval traits thus supporting the validity of the COXMM heritability estimates to forecast GWAS findings.

We next explored whether these TTE GWAS loci were novel discoveries. As GWAS of the interval phenotypes had not been previously conducted, we focused on the 65 GWAS loci-interval trait associations, which indicated 56 unique index SNPs, six of which were not identified by the corresponding case-control GWAS (**Supplementary Table 6**)^5^. Notably, only 4/65 associations were also associated (p < 5×10^−8^) with the indexing event, suggesting that index event bias was not a major driver of the interval associations (**Supplementary Table 6**). Only one locus showed an opposite direction of effect between the indexing and terminal trait, indicative of a potential collider bias (*rs429358;* **Supplementary Table 6**). In contrast, we observed that the interval associations were significant for the age-at-diagnosis of the terminal trait for 63/65 loci. We focused on two associations that were uniquely identified for the interval phenotype while having no significant association with the indexing event nor the terminal trait (**Supplementary Table 6**). The first association was *rs543040* for the progression from hypertension to type 2 diabetes trait (TTE GWAS: p=1.8×10^−9^; case-control GWAS: p=7.5×10^−7^; age-of-onset GWAS: p=5.9×10^−1^; **Fig 6**; **Supplementary Fig 6**). This association is particularly interesting as it is an intronic variant for *ABO*, a gene well linked with cardiovascular disease^27-30^. As blood group O has been shown to be protective for hypertension while being associated with poorer cardiovascular health relative to blood groups A and B, it may be that our disease progression phenotype was better powered to identify this relationship relative to an all-cause type 2 diabetes GWAS^27^. The other uniquely identified association was between *rs2754066* and the progression from hypertension to coronary artery disease trait (TTE GWAS: p=4.8×10^−8^; case-control GWAS: p=1.8×10^−6^; age-of-onset GWAS: p=6.5×10^−1^). The nearest gene, *SETD3*, was not linked with cardiovascular health through GWAS, but is known to regulate the contraction of smooth muscles with SETD3-knockout mice showing abnormal cardiac electrocardiograms and mildly decreased lean mass^31^. Taken together, our findings in the disease progression GWAS confirms that the genetic effects between the interval and terminal trait are generally shared but that interval GWAS can also identify novel associations with relevant biological mechanisms.

**Fig 6.**
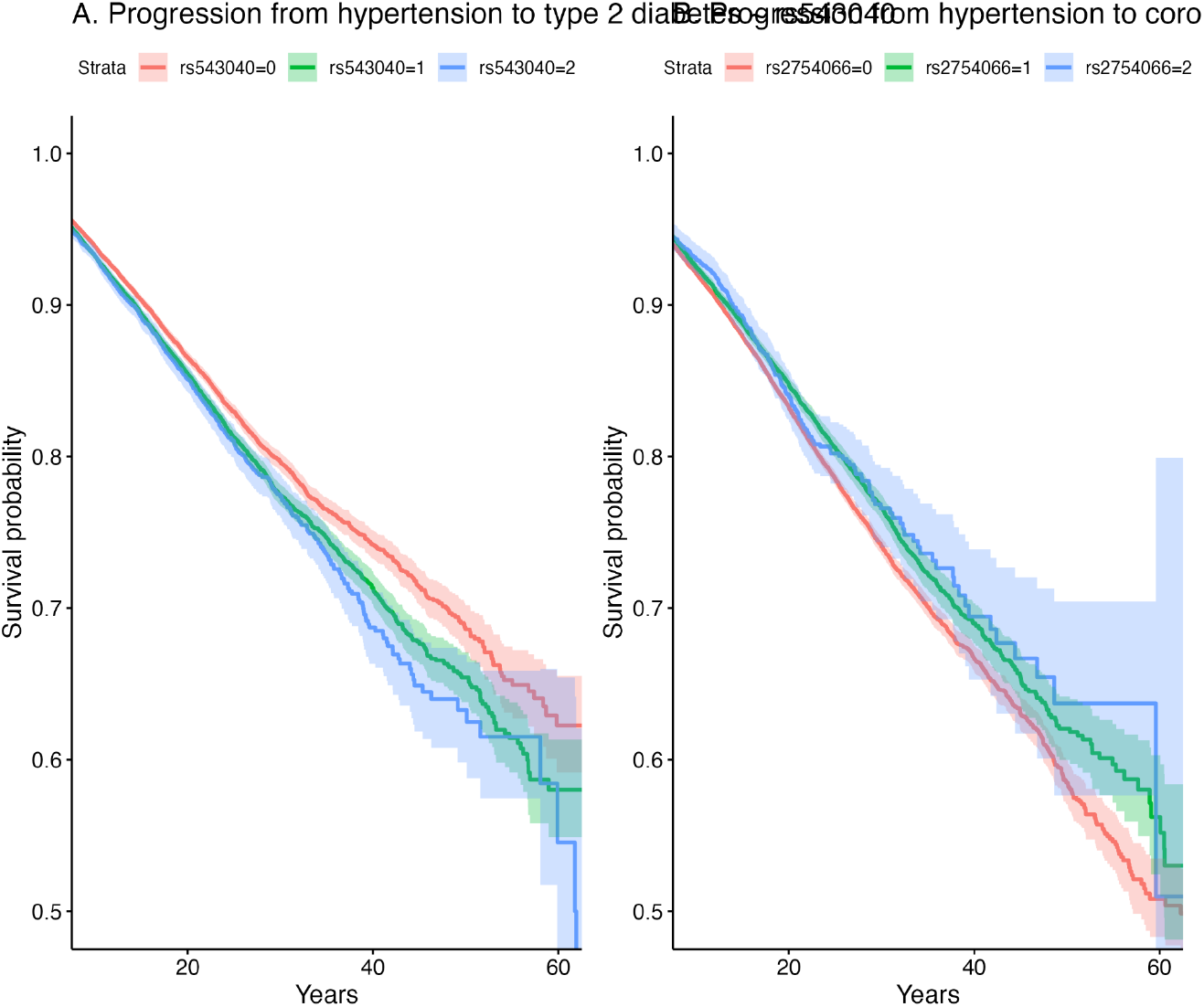
Kaplan-Meier plots for novel disease progression associations. **A**. Type 2 diabetes stratification for individuals with hypertension based on the number of copies of *rs543040*. The Kaplan-Meier plot shows the estimated survival probability over the lifecourse (or until loss to follow-up) with the plot zoomed in to events between 10 and 60 years of age. **B**. Coronary artery disease stratification for individuals with hypertension based on the number of copies of *rs2754066*. The Kaplan-Meier plot shows the estimated survival probability over the individual’s lifecourse (or until loss to follow-up) with the plot zoomed in to events between 10 and 60 years of age.

### COXMM can uncover the genetic architecture of a wide variety of traits

Finally, we applied COXMM and case-control HE-Reg to nine traits measured in the UK Biobank showcase across a variety of domains ranging from those with an adolescent onset (hayfever median age-of-onset: 20 y.o.) to late onset (dementia median age-of-onset: 75 y.o.) where we observed a strong correlation between the heritability estimates (*ρ* = 0.93, p = 2.7×10^−4^; **Fig 7A**; **Supplementary Table 3**). For 8/9 traits, the case-control HE-Reg and COXMM heritability estimates were indistinguishable, with two having a higher COXMM estimate and the remaining six having a higher case-control HE-Reg estimate though the prevailing PGM remains elusive for all eight traits (**Fig 7**; **Supplementary Table 3**). Notably, death as an outcome was not significantly heritable using either COXMM or case-control HE-Reg, indicating that low heritability for this trait cannot be explained by patient censoring or loss to follow-up. The one trait with a significantly different heritability estimate was asthma where case-control HE-Reg was higher than COXMM indicating that the LTM PGM was better aligned with this trait (Welch’s test p = 5.4×10^−6^; **Fig 7A**; **Supplementary Table 3**). As with the previously analyzed cardiovascular traits, the TTE estimates in real data were again substantially higher than the expected estimates from simulations under a pure LTM model (**Fig 7B**) and likewise for a pure TTE model (**Fig 7C**).

**Fig 7.**
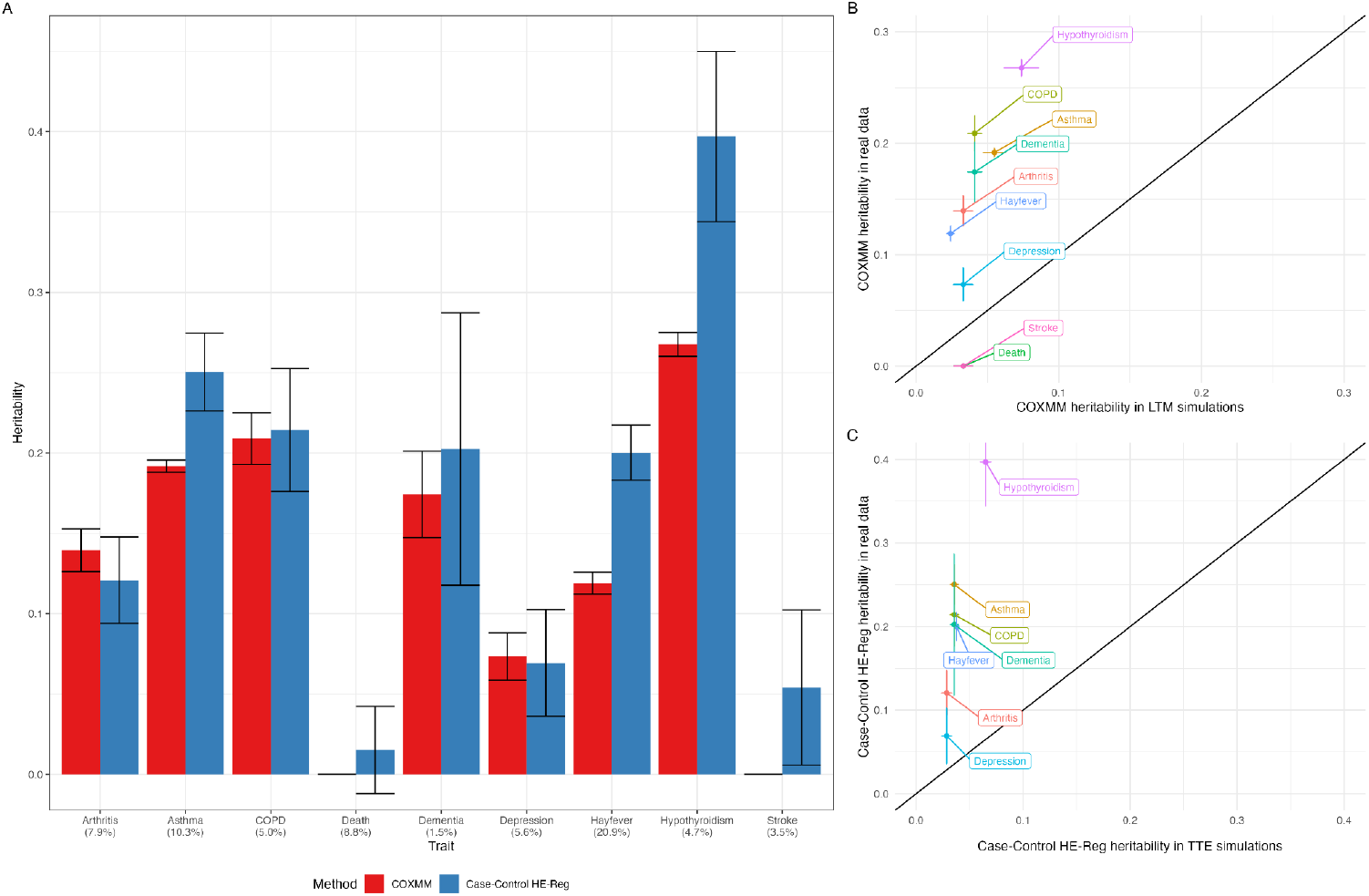
Comparison of heritability estimators using nine additional traits. **A**. We estimated the heritability of nine traits measured in the UK Biobank using two approaches. The traits are separately reported along the x-axis with a bar graph depicting the estimated heritability and standard error for each method along the y-axis: case-control HE-Reg in blue and COXMM in red. **B**. We compared the COXMM heritability estimates for the nine traits (y-axis) to the COXMM heritability estimate under the LTM PGM simulations. Each simulation’s heritability estimate for COXMM was chosen based on the ground true heritability parameter that was closest in absolute value to the Case-Control HE-Reg real data estimate and the proportion of cases/K closest in absolute value to the disease prevalence in the UK Biobank. **C**. We compared the Case-Control HE-Reg heritability estimates for the nine traits (y-axis) to the Case-Control HE-Reg heritability estimate under the TTE PGM simulations. Each simulation’s heritability estimate for Case-Control HE-Reg was chosen based on the ground true heritability parameter that was closest in absolute value to the COXMM real data estimate and the proportion of cases/K closest in absolute value to the disease prevalence in the UK Biobank. Death and Stroke were excluded due to there being no corresponding 0% heritable TTE simulations.

## Discussion

In this work we introduced COXMM, to our knowledge the only semi-parametric time-to-event (TTE) molecular heritability method offering distributional flexibility in the survival time. In prior work, TTE phenotype heritability has been estimated under accelerated failure time (AFT) models which, unlike COXMM, make parametric assumptions on the baseline hazard though are robust to violations of the proportional hazards assumption^9,10^. As AFT models do not assume the proportional hazards, they provide a fundamentally different interpretation of the genetic effect: heritability measures the proportion of variance in acceleration (or deceleration) of survival time explained by genetic factors. In contrast, Cox proportional hazard models quantify how genetics influence the instantaneous hazard at a given time point and are more widely used, particularly in GWAS^11,32-34^ In fact, semi-parametric mixed model approaches for GWAS do exist, but these methods treat the random genetic effect as a nuisance parameter and have not established its validity as a heritability estimator^6,7^. Beyond these conceptual differences, there are important performance differences worth noting. The AFT method, BayesW, was not shown to be an unbiased estimator under their own modeling assumptions and exhibited degradation in power at modest censoring levels (e.g. 40% censored/60% of cases observed)^9,10^. Another AFT method, CVC, produced unbiased estimates and demonstrated robustness to censoring down to 80% censored (20% of cases observed)^9^. COXMM, which was empirically unbiased, showed only slight deflation in heritability estimates under extreme censoring (95% censored / 5% observed cases) at higher levels of true heritability. COXMM accommodates left truncation and has been applied to such data, whereas the AFT approaches were solely focused on right-censoring events. COXXM thus differs from prior work in the modeling assumptions, parameter interpretation, and flexibility.

Here, we showed that COXMM accurately estimated heritability under various TTE simulation frameworks but underestimated the heritability when the PGM was violated, such as when the trait was simulated under the case-control PGM. We compared our method to two implementations of Haseman-Elston regression (HE-Reg): one for case-control status and the other for case-only age-of-onset. Both HE-Reg methods were downward biased under the TTE PGM. Contrasting COXMM and HE-Reg estimates can thus provide a deeper understanding of the underlying disease architecture, in addition to enabling the analysis of complex interval phenotypes.

We analyzed 34 longitudinal traits in the UK Biobank where we observed heritability estimates consistent with a mixture of a TTE and LTM PGMs. Across the traits, TTE heritability estimates were generally comparable or smaller than LTM estimates, countering the hypothesis that a substantial amount of “missing heritability” in GWAS can be explained by longitudinal modeling of trait onset or censoring. Nevertheless, TTE heritability estimates were generally higher than expected under a pure LTM model (and vice versa) suggesting that common trait architecture is a mixture of variants influencing liability and variants influencing onset. This interpretation was confirmed through the PRS and GWAS analyses, with TTE and LTM PRS frequently showing joint predictive accuracy.

Surprisingly, heritability for a terminal trait increased when excluding progression via intermediate traits, and disease progression traits generally showed significantly lower heritability than their terminal traits. Lower heritability for progression could be explained by treatments or other interventions on the index event reducing the heritability of the terminal event. Alternatively, disease progression may simply be a more stochastic event than disease incidence, as recently suggested by PRS analyses^11^. Importantly, the COXMM results were robust to age-of-onset differences and, unlike case-control HE-Reg, did not require a valid estimate of the population prevalence.

Our study has several limitations. COXMM requires a complex likelihood optimization that is computationally expensive. While we were able to analyze traits from the UK Biobank, the data had to be divided into batches (N < 30,000) and meta-analyzed. Further work is needed for improved scalability in order to systematically explore TTE traits in large biobanks. This is especially vital as COXMM requires weighted jackknifing to estimate the standard errors. Further work is also needed to understand the robustness of COXMM to censoring by competing events and/or dependent censoring, which may be particularly relevant for interval phenotypes. COXMM has also not been tested under alternative TTE frameworks such as the fully parametric AFT model or models that violate the proportional hazards assumption.

COXMM can also be extended in several ways. The inclusion of time-dependent covariates would allow adjusting for factors that change over the life-course (though this may complicate parameter interpretation if these factors are themselves heritable). Expanding COXMM to multiple variance components would also enable partitioning of TTE heritability across different genetic elements or functional annotations ^35,36^. Lastly, as access to individual level data is restricted and TTE GWAS are becoming more prevalent, there is a need for a TTE heritability method based solely on summary statistics^37^.

While we conducted an in-depth exploration of TTE traits using heritability, PRS and GWAS analyses, a number of questions remain. Our simulations indicated a clear advantage under the right PGM, and it remains unclear why the true heritability estimates were often indistinguishable for phenotypes presented in this study. If it is the case that these traits are driven by a subset of variants that act on liability and a different subset of variants that act on timing, it is of interest to distinguish these two variant classes and understand their mechanisms. The ability to estimate genetic correlation between multiple TTE traits or between early and late TTE onset would also enable identification of disease progression traits that are genetically distinct from the all-cause terminal phenotype. Ultimately, with the growing number of large-scale biobanks collecting increasingly detailed longitudinal and treatment response data, COXMM provides a systematic framework for evaluating these questions and others regarding the TTE traits. In sum, COXMM is a novel and useful tool for understanding the genetic architecture of common traits with only increasing utility as these rich datasets become available.

## Methods

### Continuous Liability Model

#### Generative Model

We now describe a number of phenotypic generative models (PGMs) starting with the standard generative model underlying quantitative traits. Let there be *N* individuals for whom we have measured *M* common SNPs and allow *X*_*N*×*M*_ to represent the standardized genotype matrix. For each individual we measure the quantitative trait *IN*_×_*I* which follows the standard Gaussian distribution.

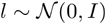

We allow *I* to be composed of two independent components: genetics, *I*_*g*_, and environment, *I*_*e*_, where *l* = *l*_*g*_ + *l*_*e*_.

Let 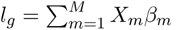 where *β M* × *I* represents the SNPs’ effect on *l* such that 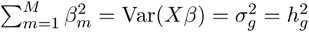 which we define as both the genetic variance component 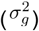 and the SNP heritability 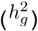 as these are equivalent under a linear model with a standardized phenotype. For now, we will refer to this parameter solely using 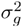 to avoid later confusion but again note the equivalence under a linear model. As a result, 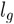 follows the Gaussian distribution:

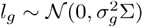

where Σ represents the Genetic Relatedness Matrix (GRM) which quantifies the genome-wide pairwise relationship between individuals. The GRM Σ is defined as:

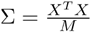

The environment component *l*_*e*_ captures the unexplained variance in the trait *l* and follows the Gaussian distribution:

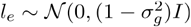

#### Estimating Heritability

There are two prevailing frameworks for estimating SNP heritability using individual-level data.

One is the Linear Mixed Model (LMM) where the vector *l*_*g*_ is treated as a random effect, and 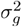 is estimated using Maximum Likelihood (ML)^38^. The other approach is the Haseman-Elston Regression (HE-Reg) which fits a linear regression to the covariance between each pair of individuals *i* and *j* ^39^.

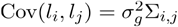

Both ML and HE-Reg have been shown to produce unbiased 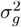 estimates for quantitative traits with ML providing a more precise estimate and HE-Reg being more scalable.

### Liability Threshold Model

#### Generative Model

We now assume the quantitative trait *I* is *unobserved* and will henceforth refer to it as the liability. Let *Y*_*N*×1_ be a binary trait representing the case-control status for each individual. We define *Y* as follows:

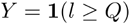

where *Q* is a scalar value representing the liability threshold defined by *K*, the population prevalence of cases. We define the threshold *Q* as:

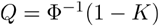

where Φ ^−1^(·) is the inverse Cumulative Density Function (CDF) of the standard Gaussian distribution.

#### Estimating Heritability

We can use ML and HE-Reg to estimate 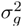 for binary traits (due to the central limit theorem). However, the 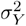 estimate is on the observed binary trait scale. In order to estimate based on 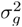 the liability scale (as defined above) for ML, a correction, *s*, is applied to 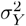 such that:

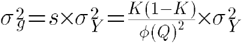

where *ϕ* (·) is the standard Gaussian density^2^. For HE-Reg, the regression requires a scaling factor *c* such that 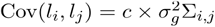 where *c* is the reciprocal of the ML correction *s*^3^. We note that sample ascertainment impacts the observed heritability estimate and therefore the correction to the liability scale. We also note the equivalence between 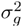 and 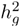 under the liability threshold model.

### Liability Induced Age-of Onset Model

#### Generative Model

While it is assumed *I* remains constant over the lifecourse, there exists a number of age-related diseases which may indicate trait prevalence varies with age^12^. We, therefore, assume that the age-of-onset *A* _*N*× 1_ is a logistic function of the liability *I* resulting in the logit *Z* _*N*× 1_:

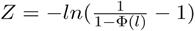

where Φ (·) is the CDF of the standard Gaussian distribution. We note, that age-of-onset, *A* is just the rescaling of *z*, such that:

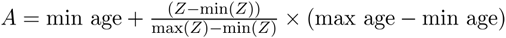

where min age and max age are the minimum and maximum age-of-onset observed in the population, respectively. Under this transformation, we assume only cases have an age-of-onset while the age-of-onset is undefined for controls.

#### Estimating Heritability

As the age-of-onset is a continuous trait, ML and HE-Reg should be well-suited for estimating 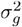 and have been used in practice after rank inverse normal transformation^23-25^. We, however, note that the estimate 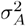 may not be on the correct scale (age-of-onset scale as opposed to the liability scale), and it excludes all controls who by definition do not have an age-of-onset.

### Time-to-Event Model

#### Generative Model

Under the liability threshold model and by extension the liability induced age-of-onset model, case-control status is an invariant function of the liability. Once case-control status is established, age-of-onset is then defined for cases; however, an alternative PGM would define the age-of-onset directly from the liability. Under this generative model and with infinite time, all individuals would become cases (i.e. develop the trait). In reality, not all cases would be observed due to censoring (e.g. death or loss to follow-up in the study) and thus be labeled as controls.

To define this more precisely, let *E*_*N ×*1_ be the time-to-event (TTE) for all individuals and *C*_*N ×i*_ be their respective censoring times. Let *D*_*N ×* 1_ be an indicator vector over all individuals denoting whether they experienced the event (i.e. were a case) or were censored (i.e. a control). More formally, for the *i* th individual d_*i*_ ^=^ **1**{e_*i*_ ≤ c_*i*_}. Let *T*= min(E, C) and be ordered such that *t*_1_ < … < *t*_*N*_.

#### Estimating Heritability

We now introduce Cox proportional hazard mixed model (COXMM) to estimate the heritability of TTE traits. Let λ(*t*)be the hazard at time *t:*

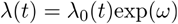

where λ_0_(*t*) is the unknown baseline hazard and *ω* is a random effect. For now, we define *ω*=*l*_*g*_*I* where *l*_*g*_ is the genetic liability and *I* is the identity matrix but will later relax this assumption to allow *ω* to capture both fixed and random effects (including the environmental component of the liability, *l*_*e*_). We note that there is no explicit modeling of error which is assumed to be captured within the unspecified baseline hazard. Furthermore, we will relax the assumption that 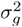 is equivalent to 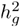 and will define *l*_*g*_ solely using the genetic variance component 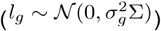.

Let *L* be the log-likelihood

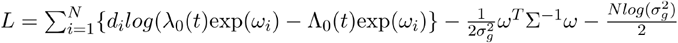

where 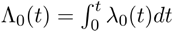 is the cumulative baseline hazard function. As λ_0_(*t*) is unknown, we define the penalized partial likelihood *L*_1_ which does not depend on the baseline hazard^13^.

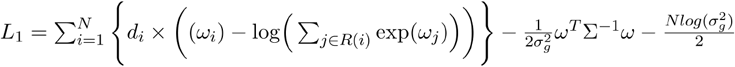

where *R*(*i*) is the set of individuals at risk at the time *t*_*i*_ when individual *i* has an event. In the above equation, 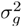 is assumed to be given and will therefore need to be estimated separately. In order to accomplish this, we define the integrated marginal likelihood *L*_2_ using the Laplace approximation^13,14^:

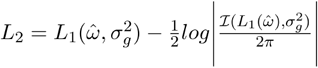

where *ℐ* is the Fisher information matrix with respect to the random effect *ω*. In order to produce the estimate 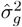, we iterate between the two equations (*L*_1_ and L_2_).

We compute the score function and Fisher Information matrix with respect to *ω* for *L*_1_ and note that any ties in *T* are addressed using Breslow’s approximation^17^. We use Newton-Raphson (NR) to update *ω* until convergence. We note the complexity is 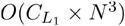 due to performing Cholesky decomposition for each update where 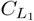 is the number of updates but note that 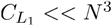 particularly in biobank scale datasets.

As 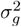 is a scalar, we use a derivative free optimization (BOBYQA) and treat *L*_*2*_ as a marginal likelihood of 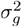^13,40^. Therefore, in each iteration, we optimize *L*_1_ as defined above using NR and then evaluate the 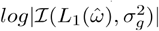 which requires a Cholesky decomposition. As a result the complexity of this process is 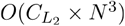 where 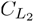 is the number of iterations. We note that empirically, 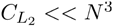 especially in biobank scale datasets. Thus resulting in an overall cost of *O*(*N*^3^).

We note the baseline hazard λ_0_(*t*) is by definition undefined and therefore the error is not directly modeled in this semiparametric model. As a result, COXMM would likely capture much of the environmental effects within the unspecified baseline hazard.

### Definition of variance in COX proportional hazard Mixed Model

As stated above, linear models have the following well known property that if 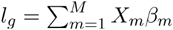 where *x* is a standardized genotype matrix of the *M* SNPs, β_*m*_ is the effect size of each SNP, and *l* = *l*_*g*_ + *l*_*e*_, then 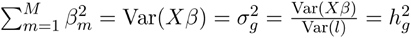. This property allows the parameter 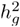 to represent the proportion of variance explained as well as the sum-squared effects/genetic variance component.

Unfortunately, the Cox proportional hazard model does not have a phenotypic variance defined 2 as it relies only on the relative ordering of events. This does not mean that the parameter 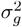 is not informative as it still reflects the sum-of-squared effects and captures the genetic variance component of the liability. It does not, however, represent the proportion of variance explained in the classical linear model sense.

Fortunately, previous work has shown that under our generative model (also known as a semiparametric Cox frailty model), heritability can be measured on the log frailty scale^18^. Let 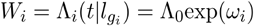 be the conditional integrated hazard function for individual *i* which provides a measure of the total accumulated risk of an event. As shown in the work of Korsgaard et. al, we will define ϵ _*i*_ as the log of the accumulated risk for individual *i* and note it follows the extreme value distribution:

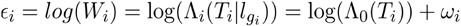

Through rearrangement of the terms, it can be shown that this equation is equivalent to a linear model on the log cumulative baseline hazard scale (log (Λ_0_(*T*_*i*_))). The unconditional variance of log (Λ_0_(*T*_*i*_)) is 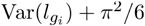_;_ therefore the definition of heritability on the log cumulative baseline hazard scale (log (Λ_0_)) is as follows:

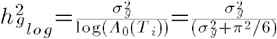

### Estimating Heritability with Fixed Effects present

#### Continuous Liability, Liability Threshold Model, and Liability Induced Age-of-Onset Model

Let *V*_*N ×P*_ be the set of *p* covariates with the effect sizes γ_*P* ×1_ such that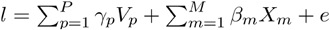. For the continuous trait *l* (*unobserved* liability), the heritability estimate *ĥ*^*2*^ produced using ML is biased due to the presence of fixed effects. In order to correct this, REstricted Maximum Likelihood (REML) is applied to a transformation of the data such that the fixed effects have no impact on the heritability estimate *ĥ*^*2*^.

As the LMM is used to estimate 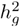 under the liability threshold model (case-control status) and liability induced age-of-onset, REML is the appropriate method when covariates are present in these instances as well. We, however, note that previous work has shown that for *N > 2*,500 REML produced downward biased heritability estimates for case-control traits while HE-Reg remained unbiased ^3^.

#### Time-to-Event Model

While substantial evidence has shown that REML is necessary for LMM, there is currently no evidence supporting the superiority of REML over ML for Cox proportional hazard mixed models; therefore, COXMM is only implemented as a ML method. We note that the fixed effects are explicitly modeled in *ω* such that *ω* = *γV*+*l*_*g*_ and therefore their effect sizes are jointly estimated with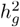. Empirically we show the presence of fixed effects have a negligible effect on the heritability estimate (see **Fig. 2**).

### Simulations

#### General Parameters

For each set of simulations, we created 10,000 unrelated individuals with 2,500 independent SNPs. All SNPs have a minor allele frequency (MAF) > 0.05. We considered up to eight values for the variance component 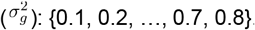. We assumed each SNP had an effect size drawn from the infinitesimal model. We used seven values for the proportion of observed events/cases (*K*): {0.01, 0.05, 0.1, 0.2, 0.4, 0.65, 0.72}. We generated 50 replicates for each set of analyses and reported the mean heritability estimate and its standard error. We set the age-of-onset and age-at-censoring to be between 30 and 70, but note that the ages are arbitrary and simply a rescaling factor.

#### Case-Control Generative Model

We simulated case-control status according to the liability threshold model and set the age-of-onset according to a liability induced age-of-onset model using a logit function (see above). As COXMM expected a censoring age for controls, we simulated the censoring age in an identical fashion but instead of using the liability, we generated an independent standard Gaussian to inform the age-at-censoring.

#### Time-to-Event Generative Model

We generated TTE traits with a uniform baseline hazard and set the proportional hazard using a Weibull distribution with 1 as the shape parameter and the genetic liability as the scale parameter. We censored the later quantile of events to achieve the appropriate proportion of observed events/cases in a manner that reflects cohort censoring.

In addition to the above simulations, we also separately adjusted the simulation framework as follows:

1. Simulated a Weibull distribution with 3 as the shape parameter (scale parameter still *l*_*g*_)
2. Added environment effect *l*_*e*_ to the scale parameter such that the total variance of *ω* = 1 = 1
3. Added 2 fixed effect standard normal variables contributing variance = 0.01 to the scale parameter
4. Added 2 fixed effect standard normal variables contributing variance = 0.5 to the scale parameter
5. Generated a censoring event using the exponential distribution with a rate resulting in *K =* 0.65
6. Generated the censoring event using the exponential distribution with a rate resulting in *K =* 0.72

#### Analysis Software

When we treated the simulated phenotypes (regardless of PGM) as a TTE trait, we ran the heritability analysis using COXMM (https://github.com/koditaraszka/COXMM). For case-control phenotypes, we used PCGC as implemented in LDAK but note we assumed all SNPs contributed equally^1,3,4^. We also assumed PCGC was given the true population proportion of cases in all simulations. For age-of-onset phenotypes, we used HE-Reg also implemented in LDAK but again assumed all SNPs contributed equally. We used rank inverse normal transformation (RINT) to standardize the age-of-onset (which reflected our data processing steps in the real data analyses).

### UK Biobank

#### Phenotype Generation

The UK Biobank data was obtained under application 33217. From which we generated seven cardiovascular traits using the ukpheno R package^26^. This software package used a conglomerate of data sources from the UK Biobank (e.g. self-reported data, ICD codes, medications, hospital records, etc.). We further generated 18 interval phenotypes using the interval between the age-of-onset for three traits: type 2 diabetes, hypertension, and hyperlipemia and the remaining traits. For the analyses, we restricted to 276,194 unrelated white Britons and processed SNPs such that the MAF≥ 0.01, missing genotype rate ≤ 0.01, and Hardy-Weinberg p-value ≥ 1×10^−7^. We also included sex and the top 10 in-sample principal components as covariates for all analyses. For TTE traits, we set the censoring age to the first occurrence of one of the following: date lost to follow-up, death, or the last data collection date for each center. For age-of-onset analyses, we applied a RINT to the data (**Supplementary Table 3**).

#### Heritability Analyses

For the heritability analyses, we included 531,269 genotyped SNPs in the GRM. Due to computational limitations with COXMM, we split the data into ten approximately equal groups and meta-analyzed across the groups for each method. We note that each of these subsets contained approximately 30,000 individuals and was analyzed using up to 64G of RAM. Furthermore, previous work has indicated that Cox proportional-hazard mixed models underestimated the standard error for variance components, so we computed the standard errors for COXMM using a weighted block jackknife ^15,16^.

#### PRS Analyses

For the PRS analyses, we restricted to 90% of the cohort (N = 248,574) and ran the case-control GWAS using logistic regression and the TTE GWAS using SPACox ^5^. We then LD pruned the SNPs (M = 954,195) and projected their estimated z-scores onto the held-out sample for 27,620 individuals. After generating a TTE PRS and case-control PRS and standardizing both, we fit four Cox proportional hazard mixed models: the PRSs jointly fit, each PRS alone, and a null model. We then separately fit these four models in a logistic regression. We included the standard covariates in all models.

#### GWAS Analyses

For the GWAS analyses, we ran SPACox for the TTE trait, logistic regression for case-control status, and a linear regression for the age-of-onset^5^. As SPACox was not a mixed model, none of the analyses used a mixed model to enable a fair comparison. We reported results on the 4,871,483 imputed SNPs which passed QC (see above).

## Data Availability

All software and scripts for data generation/figure generation is available online at https://github.com/koditaraszka/COXMM/

## Supplementary Figures

**Supplementary Fig 1.**
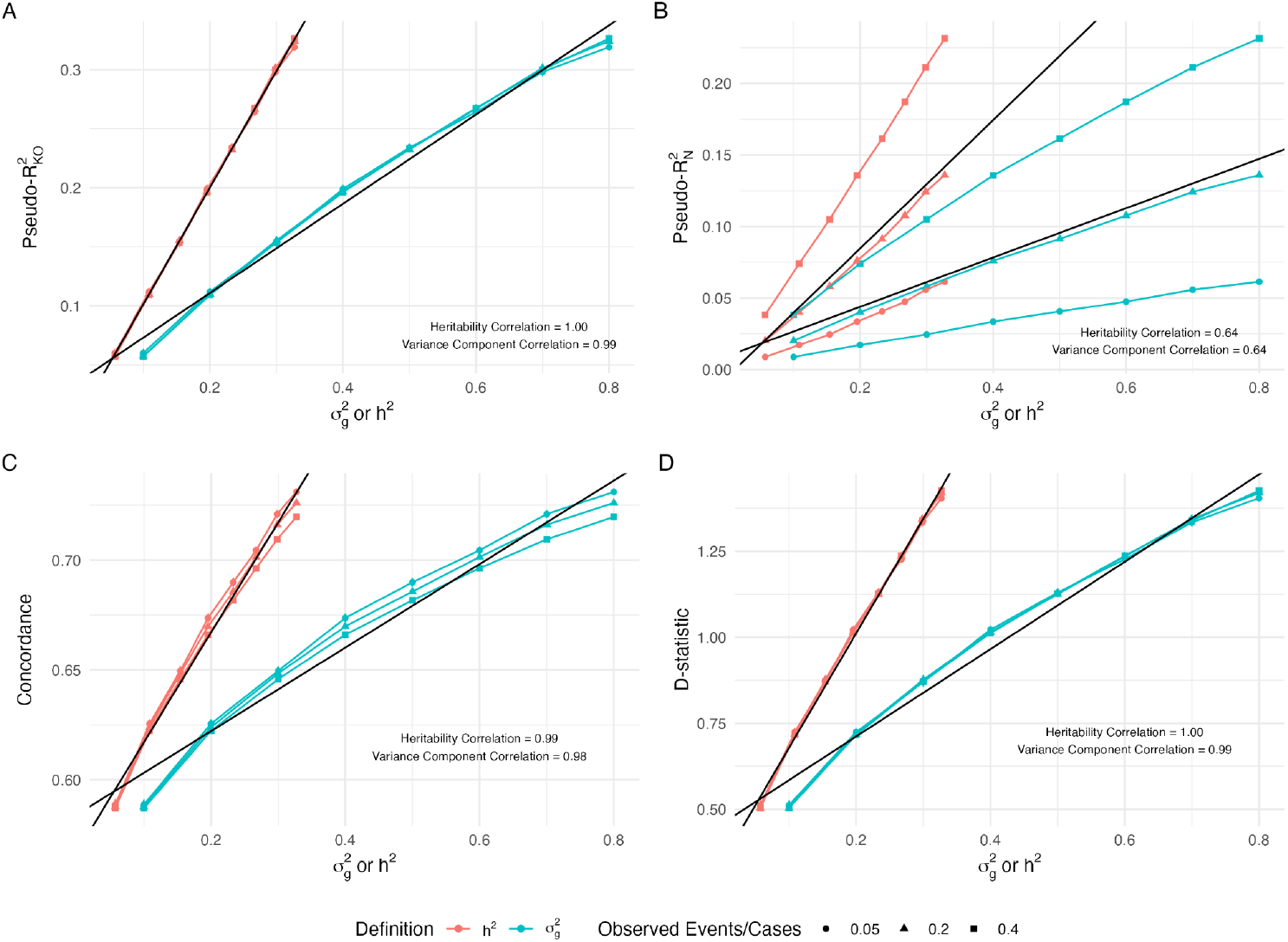
COXMM performance under various phenotypic generative models (PGMs) including low prevalence of events. We simulated 50 traits under the time-to-event (TTE) PGM using a Weibull distribution with the shape parameter set to 1 and the scale parameter accounting for the genetic liability. This was repeated for each value of 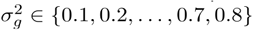 and proportion of observed events/cases (*K* ∈ {0.05,0.2,0.4}). Within each panel, the x-axis corresponded to the true variance component 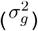 or the corresponding transformed heritability 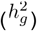. The y-axis reflected the average model fit metric for the heritability in red and for the variance component in blue with a black line reflecting the linear model fit. **A**. The y-axis is the pseudo-R^2^ defined by Kent and O’Quigley. **B**. The y-axis is the pseudo-R^2^ defined by Nagelkerke. **C**. The y-axis is the concordance. **D**. The y-axis is the Royston and Sauerbrei d-statistic.

**Supplementary Fig 2.**
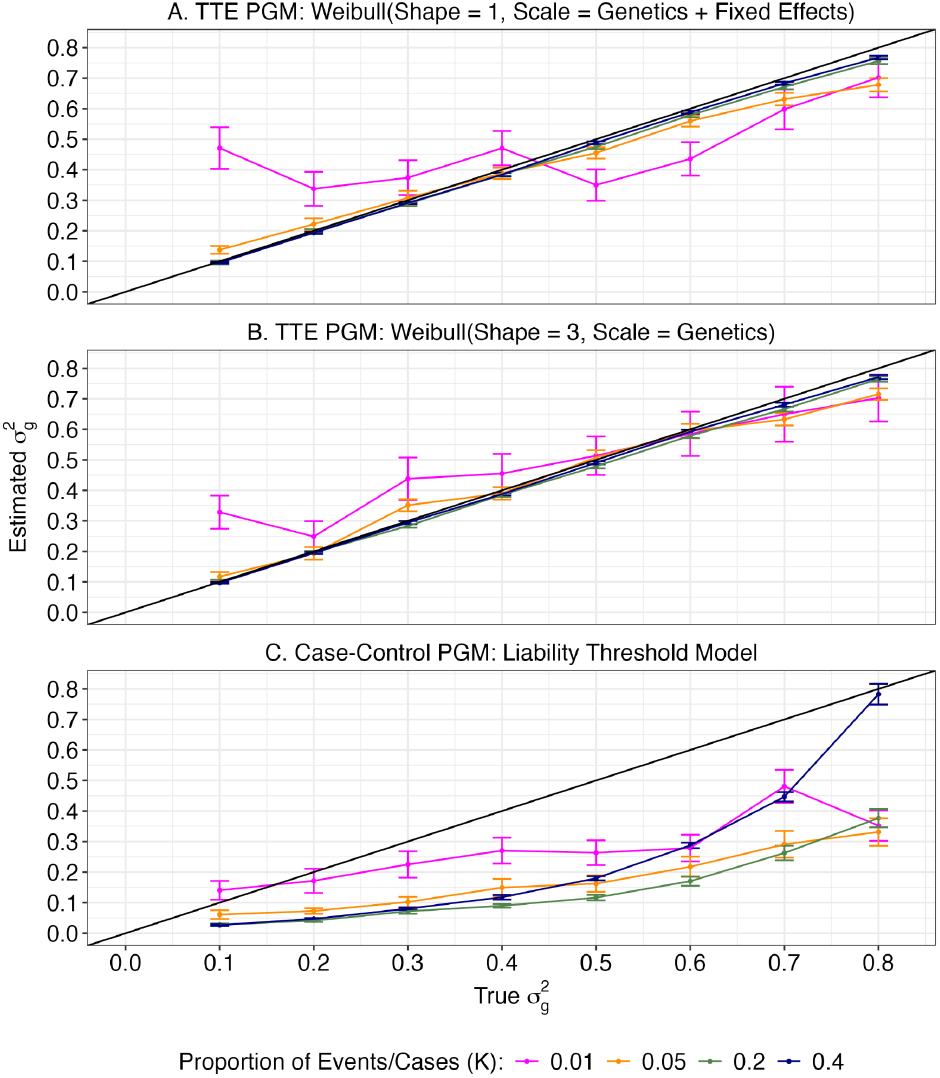
COXMM performance under various phenotypic generative models (PGMs) including low prevalence of events. In each panel, we simulated a different phenotypic generative model (PGM) across heritability levels 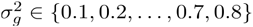 and proportion of observed events/cases (*K* ∈ {0.01,0.05,0.2,0.4}). Within each panel, the x-axis corresponded to the true heritability and the y-axis reflected the estimated heritability with a black line indicating the ground truth. **A**. We simulated 50 traits under the time-to-event (TTE) PGM using a Weibull distribution with the shape parameter set to 1 and the scale parameter accounting for the genetic liability and two fixed effect variables with the total fixed effect variance set to 0.5. **B**. We simulated 50 traits under the TTE PGM using a Weibull distribution with a shape parameter of 1 and the scale parameter only based on the genetic liability. **C**. We simulated 50 traits under the case-control PGM using the classic LTM.

**Supplementary Fig 3.**
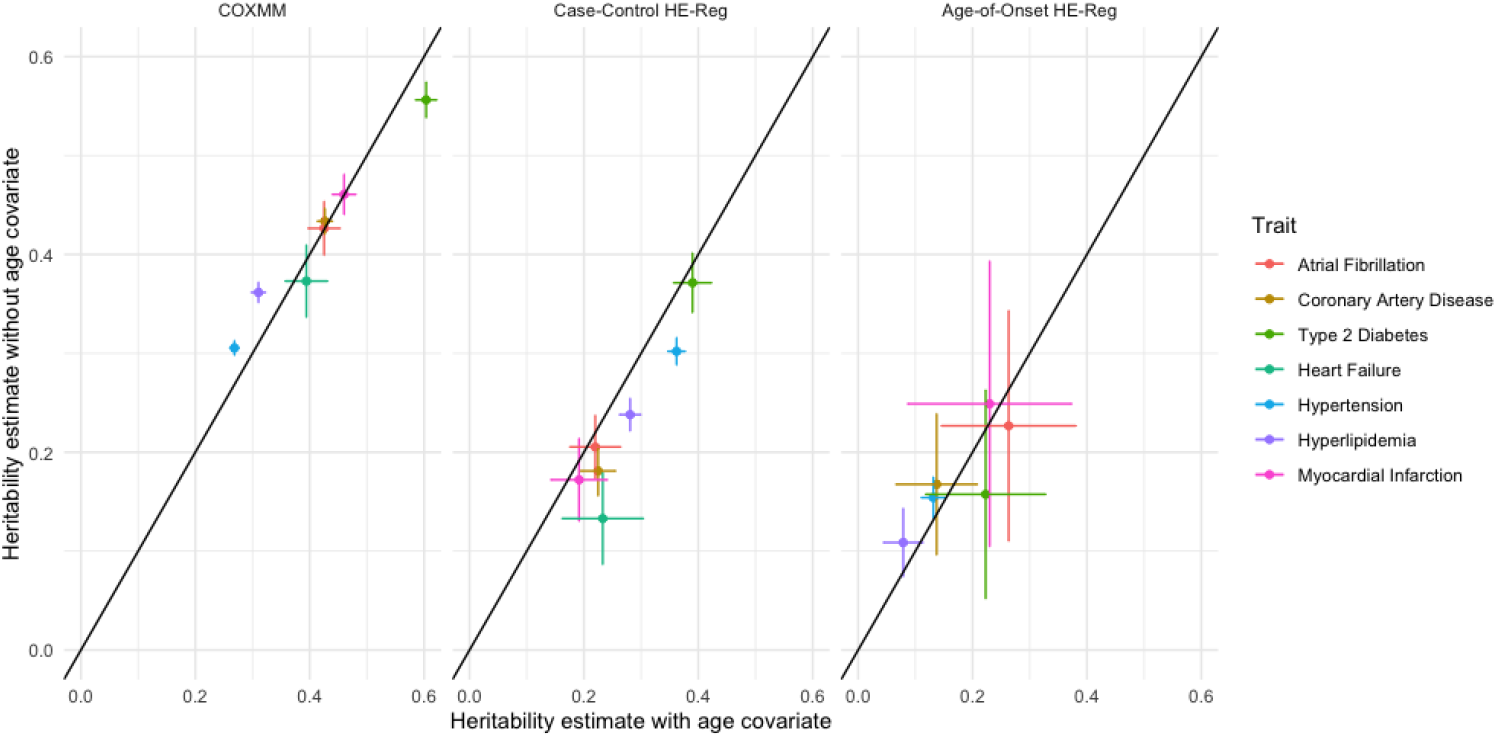
COXMM is not biased by the age-at-enrollment covariate. We compared the heritability estimates generated with and without the inclusion of the age-at-enrollment covariate for the seven age-at-diagnosis traits. We report the estimate with the age covariate on the x-axis and without the covariate on the y-axis for COXMM, case-control HE-Reg and age-of-onset HE-Reg in separate panels.

**Supplementary Fig 4.**
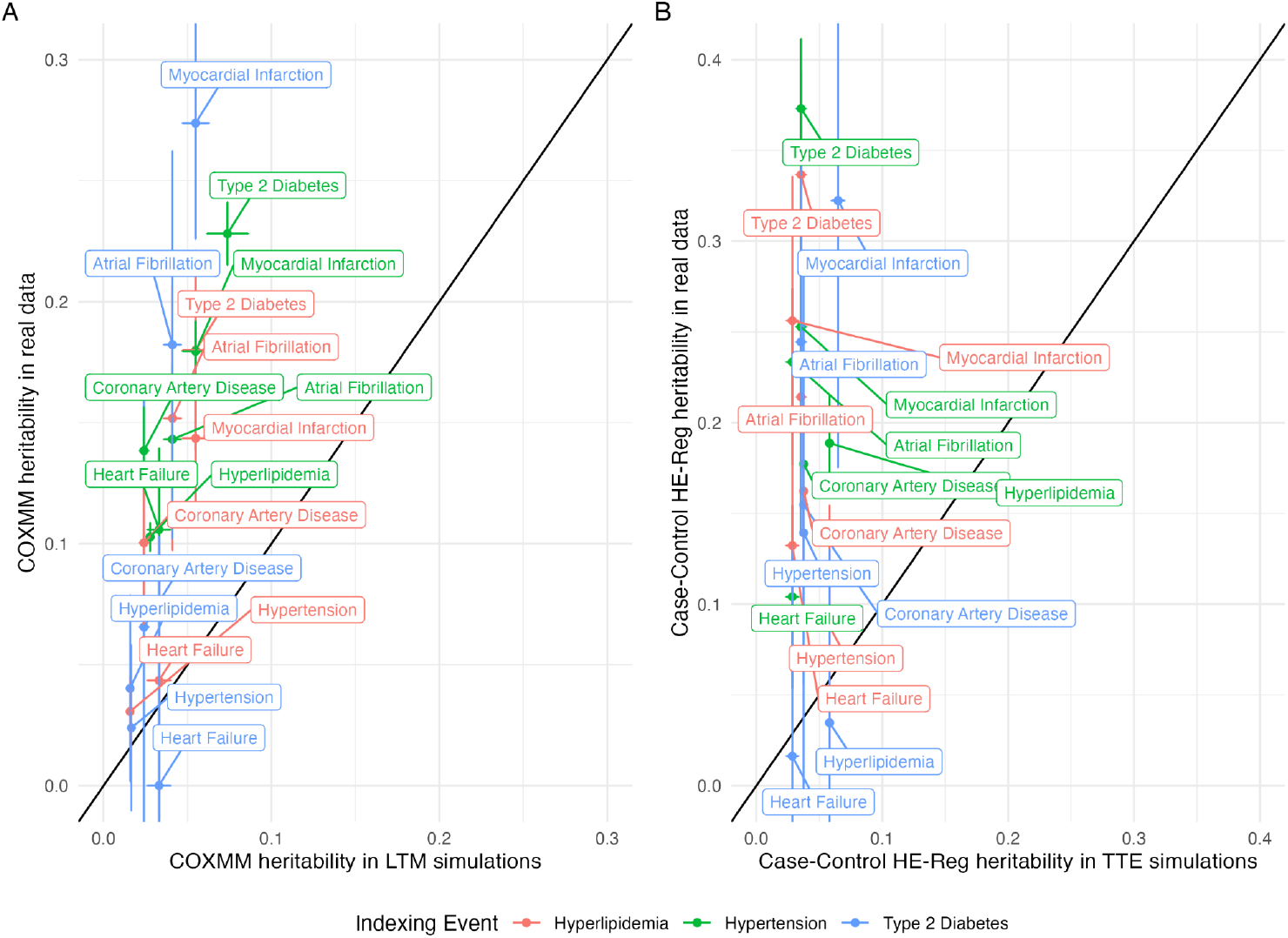
Comparison of real data heritability estimates and expected heritability estimates based on the PGM. **A**. We compared the COXMM heritability estimates for the 18 interval traits (y-axis) to the COXMM heritability estimate under the LTM PGM simulations. Each simulation’s heritability estimate for COXMM was chosen based on the ground true heritability parameter that was closest in absolute value to the Case-Control HE-Reg real data estimate and the proportion of cases/K closest in absolute value to the disease prevalence in the UK Biobank. The data points were color coded based on the indexing event and annotated with the terminal event. **B**. We compared the Case-Control HE-Reg heritability estimates for the 18 interval traits (y-axis) to the Case-Control HE-Reg heritability estimate under the TTE PGM simulations. Each simulation’s heritability estimate for Case-Control HE-Reg was chosen based on the ground true heritability parameter that was closest in absolute value to the COXMM real data estimate and the proportion of cases/K closest in absolute value to the disease prevalence in the UK Biobank. The data points were color coded based on the indexing event and annotated with the terminal event.

**Supplementary Fig 5.**
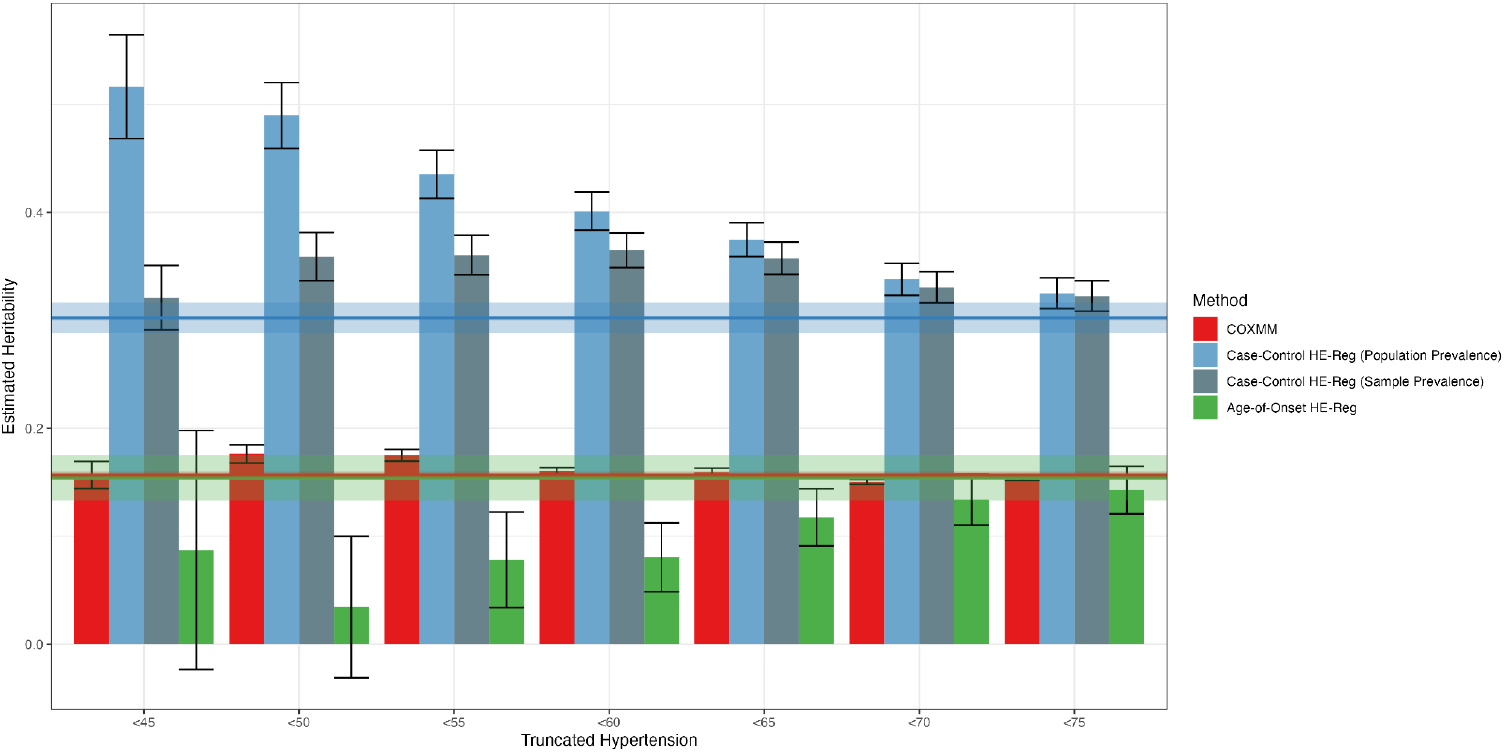
COXMM is robust to phenotype truncation. We estimated the heritability of the age-at-diagnosis for hypertension measured in the UK Biobank using three approaches case-control HE-Reg in blue, case-only age-of-onset HE-Reg in green, and COXMM in red and report the cohort level estimate and its standard error using a horizontal line. We truncated the age-at-diagnosis by censoring everyone with a data point past the cutoff age. For case-control HE-Reg, we transform to the liability scale using the sampling prevalence of cases at the truncated age and the full population prevalence of cases (0.4).

**Supplementary Fig 6.**
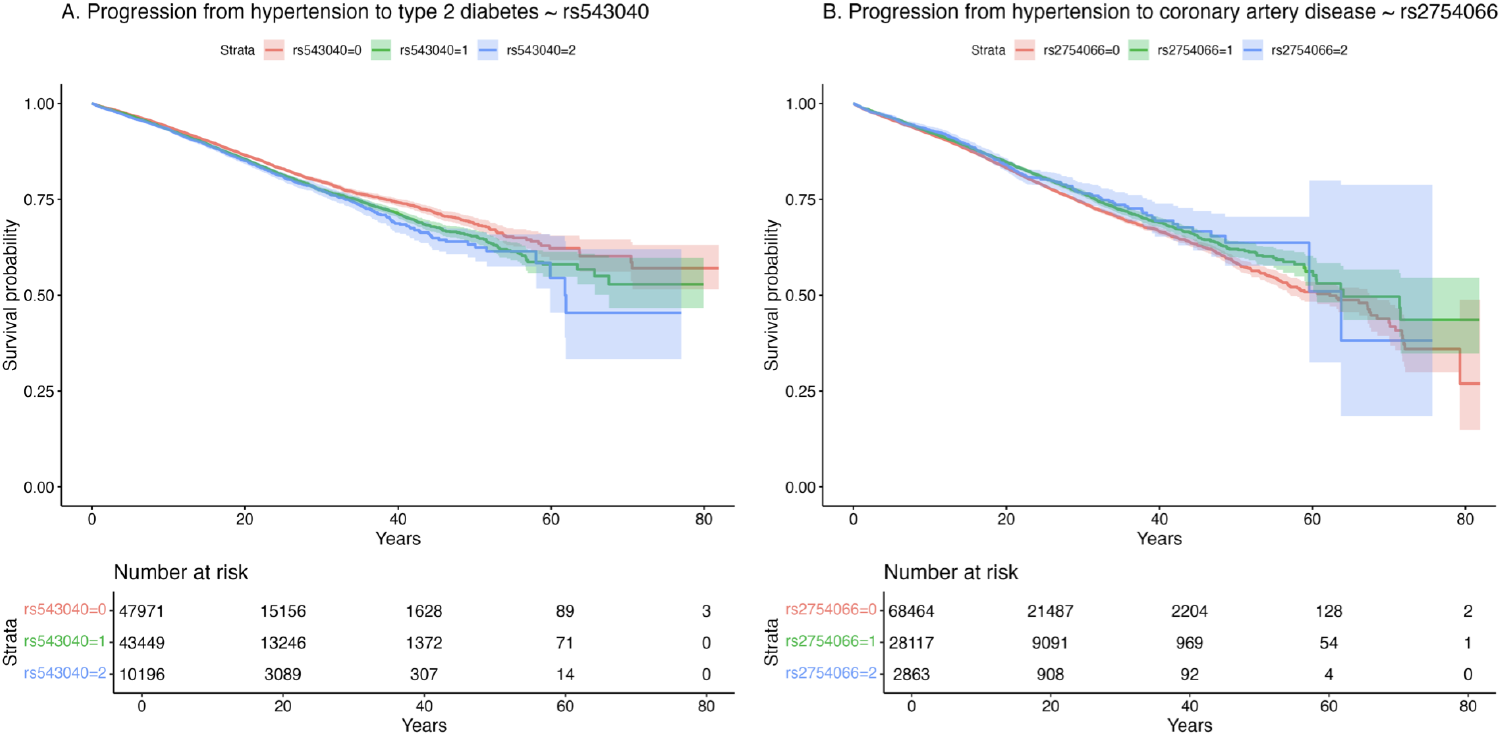
Kaplan-Meier plots for novel disease progression associations. **A**. Type 2 diabetes stratification for individuals with hypertension based on the number of copies of *rs543040*. The Kaplan-Meier plot shows the estimated survival probability over the lifecourse (or until loss to follow-up). **B**. Coronary artery disease stratification for individuals with hypertension based on the number of copies of *rs2754066*. The Kaplan-Meier plot shows the estimated survival probability over the individual’s lifecourse (or until loss to follow-up).

